# Estimating the within-person change in dental service access measures during the COVID-19 Pandemic

**DOI:** 10.1101/2022.06.08.22276174

**Authors:** Jason Semprini

## Abstract

**Background:** American adults delay dental care more than any other healthcare service. Unfortunately, the COVID-19 pandemic may have stalled efforts to address dental service delays. Early evidence has suggested substantial declines in dental service visits in the early phase of the pandemic, however our study is among the first to measure within-person changes from 2019 to 2020 and conduct subgroup analyses to examine if changing dental patterns were mediated by exposure to the pandemic, risk of adverse COVID-19 outcomes, or dental insurance.

**Methods:** We analyzed a National Health Interview Survey panel of individuals initially surveyed in 2019, with subsequent follow up in 2020. The outcomes included dental service access measures and the interval of a most recent dental visit. By constructing a probability weighted linear regression model with fixed-effects, we estimated the average within-person change from 2019 to 2020. Robust standard errors were clustered within each respondent.

**Results:** Overall, adults in 2020 were 4.6%-points less likely to visit the dentist compared to 2019 (p < 0.001). Significantly higher declines were found in Northeast/West regions compared to Midwest/South. We find no evidence that declining dental services in 2020 were associated with more chronic diseases, older age, or lack of dental insurance coverage. Adults did not report more financial or non-financial access barriers to dental care in 2020 compared to 2019.

**Conclusions:** The long-term effects of the COVID-19 pandemic on delayed dental care warrants continued monitoring as policymakers aim to mitigate the pandemic’s negative consequences on oral health equity.

## Introduction

American adults delay dental care more than any other healthcare service [1]. Higher rates of delayed care are associated with poor health and socioeconomic status [2-4]. Adults who do not regularly (at least annually) visit a dentist have been found to be at higher risk of developing caries, Periodontitis, and mouth pain, which are among the most prevalent chronic conditions in the country [5-6]. Further delaying treatment for chronic oral health conditions can lead to invasive and expensive treatment in the future. Left untreated, chronic oral health conditions may eventually result in tooth loss and low functional dental status [7-9].

Delaying dental services is a major public health issue. Unfortunately, the COVID-19 pandemic may have stalled efforts to address dental service delays [10, 11]. International reports suggested dental visits declined substantially early in the pandemic [12, 13]. Studies on dental visits in the U.S. found lower utilization in 2020 for children and adults [14-17]. These U.S. studies, however, rely on cell-phone tracking data and cross-sectional surveys that may fail to account for unobserved individual level factors associated with dental service use.

### New Contributions

Measuring a change within a single person over time is typically expensive and inaccessible for most population-based health services research. However, the within-subject research design can mitigate numerous threats to internal validity and requires much fewer statistical and causal inference assumptions. We are among the first to estimate the within-person changes in dental service access measures by analyzing a novel, population-based cohort dataset. We are also among the first to identify changing patterns of dental service access and utilization measures by subgroups associated with exposure to COVID-19 cases, and risk of adverse COVID-19 outcomes, and propensity to visit a dentist. Finally, we explore the mechanisms which may have influenced decisions to delay dental visits by not only using utilization measures, but also analyzing self-reported measures indicating if the respondent delayed dental care due to cost and if the individual was unable to obtain dental care. Understanding which adults delayed dental care, and why, can help shape policy responses to increase access to dental services and mitigate oral health disparities resulting from delayed dental treatment.

### Conceptual Framework

Emerging research affirms that adult dental services declined immediately following the pandemic [14, 15] However, even in areas where dental services remained available or returned to full capacity, certain factors may have mediated the relationship between the COVID-19 pandemic and decisions to visit the dentist. One factor is the exposure to the pandemic. The pandemic affected everyone, but consider the difference in exposure to the pandemic for two adults: an adult in New York City (Northeast, Large Metro) compared to an adult in rural South Dakota (Midwest, Nonmetro) [18]. These two adults had vastly different exposures to COVID-19 case rates, lockdown policies, and social distancing norms. If exposure to the pandemic is a factor for changing dental service utilization, we would expect greater changes in dental visits for adults living in metro areas and in the Northeast/West regions, compared to changes in adults living in nonmetro areas and Midwest/South regions.

Another potential factor mediating the relationship between the COVID-19 pandemic and dental services is the risk of acquiring COVID-19 at a dental visit. Social distancing behaviors are more beneficial to adults facing higher likelihood and consequences of adverse outcomes following a COVID-19 infection. If risk is a determinant of changing dental visits during 2020, we would expect to observe a larger decline in dental service utilization for high-risk (older, sicker) populations. We test this hypothesis by identifying differences in changing dental services by age and disease status.

A final component of our conceptual model is dental insurance coverage. The decision to obtain private dental insurance coverage is clearly endogenous with the decision to seek dental services in the future [19]. However, the decision to obtain dental insurance is independent of the COVID-19 pandemic (we explicitly test this assumption), as the enrollment decisions were likely made before March 2020. Whether an adult obtained dental insurance prior to the COVID-19 pandemic can serve as a measure of their propensity to visit a dentist in the upcoming year. We predict that the COVID-19 pandemic did not uniformly impact dental services behavior and expect a larger change from 2019 to 2020 for adults without dental insurance coverage in 2020.

## Methodology & Materials

### Data and Sample

We analyzed a two-year panel of individuals initially surveyed in 2019, with subsequent follow up in 2020. This nationally representative, population-based data came from the National Health Interview Survey (NHIS) for years 2019-2020, and was extracted from the Integrated Public-Use Micro Data Series (IPUMS) [20, 21].

### Variables

The primary outcome of interest is whether the respondent visited the dentist in the past year. This binary measure is derived from a survey question asking about the interval since the last dental visit. Additional outcome variables aim to measure unmet dental needs, one being a binary variable indicating if the respondent delayed dental care in the past year due to cost and another indicating if the respondent was unable to get dental care in the past year. We also include private dental coverage as an additional binary outcome variable.

### Research Design & Analytical Strategy

Under a panel design, we estimated the average effect of the pandemic on dental visits with a simple pre-post, within-person design: E[Dental Visit < 1 Year| POST=1, ID = i] – E[Dental Visit < 1 Year| POST = 0, ID=i], where ID is the individual i and POST = 1 indicates the year is 2020.. We then constructed subgroup analyses by stratifying samples into categories based region (NE, W, S, MW), metro status (large metro, fringe metro, small metro, nonmetro), chronic disease status (0, 1+), co-occurring disease status (no occurring conditions, co-occurring conditions), and age (18-45, 45-64, 65-74, 75-84). A final subgroup analyses tests for differences between adults with and without private dental insurance coverage.

All analyses were constructed as a linear probability regression model which included within-person fixed-effects. For inference and determining statistical significance, we estimated standard errors robust to heteroskedasticity and serial autocorrelation, clustered within each individual. All analyses incorporated NHIS supplied cohort sampling weights.

## Results

Our sample includes 10,415 adults, which were surveyed in both 2019 and 2020. Supplemental tables 1 and 2 report the sample characteristics and baseline (2019) outcomes for the full sample and subgroups. Table 1 reports the average within-person change in dental service outcomes for the full sample. Table 2 reports the subgroup estimates. The average change in the proportion of adults visiting the dentist in the past year is shown in Figure 1 (region, metro), Figure 2 (chronic disease, co-occurring condition), and Figure 3 (age, dental insurance coverage).

**Table 1:** Average Within-Person Changes in Dental Service Outcomes: 2019 to 2020, full sample.

|  | Estimate | (se) |
| --- | --- | --- |
| *<u>Access Measures</u>* |  |  |
| Delayed Dental Care Due to Cost | -0.013* | (0.006) |
| Unable to Obtain Dental Care | -0.021*** | (0.006) |
| *<u>Utilization Measures</u>* |  |  |
| Last dental visit <1 year | -0.046*** | (0.007) |
| Last dental visit 1-2 years | 0.041*** | (0.007) |
| Last dental visit 2-3 years | 0.004 | (0.005) |
| Last dental visit 3-5 years | -0.001 | (0.004) |
| Last dental visit 5+ years | 0.002 | (0.004) |
| Never visited dentist | -0.001 | (0.001) |
| *<u>Insurance Measures</u>* |  |  |
| Enrolled in Dental Insurance | -0.000 | (0.006) |

Table 1 reports the coefficients from our primary analysis, estimating the within-person change in dental outcomes in 2020 compared to 2019. All analyses were estimated by linear probability regression model with individual fixed-effects. Robust standard errors were clustered at the individual level and reported in parentheses.
\* $p < 0.05$ , \*\* $p < 0.01$ , \*\*\* $p < 0.001$ .

**Table 2:** Average Within-person changes in Dental Service Outcomes: 2019 to 2020, by subgroups.

|  | Delayed Dental<br>Care Due to Cost |  | Unable to Obtain<br>Dental Care |  | Last dental visit<br><1 year |  |  |
| --- | --- | --- | --- | --- | --- | --- | --- |
| Group | Estimate | (se) | Estimate | (se) | Estimate | (se) | N |
| <u>Region</u> |  |  |  |  |  |  |  |
| Northeast | -0.013 | (0.014) | -0.039** | (0.014) | -0.061*** | (0.016) | 1,778 |
| Midwest | -0.001 | (0.011) | -0.008 | (0.011) | -0.018 | (0.013) | 2,428 |
| South | -0.018 | (0.011) | -0.022* | (0.010) | -0.039*** | (0.011) | 3,538 |
| West | -0.017 | (0.012) | -0.017 | (0.012) | -0.069*** | (0.015) | 2,614 |
| <u>Metro</u> |  |  |  |  |  |  |  |
| Large Metro | -0.001 | (0.011) | -0.026* | (0.011) | -0.045*** | (0.013) | 3,075 |
| Fringe Metro | -0.022 | (0.012) | -0.019 | (0.012) | -0.039* | (0.016) | 2,278 |
| Small Metro | -0.011 | (0.011) | -0.019 | (0.011) | -0.054*** | (0.011) | 3,285 |
| Nonmetro | -0.029* | (0.014) | -0.017 | (0.012) | -0.041** | (0.016) | 1,719 |
| <u>Chronic Disease</u> |  |  |  |  |  |  |  |
| <1 Diagnoses | 0.001 | (0.010) | -0.004 | (0.009) | -0.061*** | (0.012) | 3,782 |
| 1+ Diagnoses | -0.025** | (0.008) | -0.033*** | (0.008) | -0.040*** | (0.009) | 6,575 |
| <u>Co-Occurring</u><br><u>Conditions</u> |  |  |  |  |  |  |  |
| <2 Diagnoses | -0.005 | (0.007) | -0.012 | (0.007) | -0.052*** | (0.009) | 6,622 |
| 2+ Diagnoses | -0.037** | (0.012) | -0.050*** | (0.011) | -0.032** | (0.011) | 3,735 |
| <u>Age</u> |  |  |  |  |  |  |  |
| <45 Years | 0.001 | (0.011) | -0.002 | (0.011) | -0.064*** | (0.014) | 2,610 |
| 45-64 Years | -0.018 | (0.009) | -0.029*** | (0.009) | -0.037*** | (0.010) | 4,227 |
| 65-74 Years | -0.034* | (0.014) | -0.030* | (0.012) | -0.039** | (0.014) | 2,055 |
| 75+ Years | -0.014 | (0.017) | -0.035* | (0.016) | -0.042* | (0.019) | 1,465 |
| <u>Dental Insurance</u> |  |  |  |  |  |  |  |
| No Dental Insurance | -0.014 | (0.008) | -0.021** | (0.008) | -0.050*** | (0.009) | 8,034 |
| Yes Dental Insurance | -0.019 | (0.013) | -0.016 | (0.010) | -0.042* | (0.019) | 2,264 |

Table 2 reports the coefficients from our primary analysis, estimating the within-person change in dental outcomes in 2020 compared to 2019. Each model was separately analyzed for each subgroup. All analyses were estimated by linear probability regression model with individual fixed-effects. Robust standard errors were clustered at the individual level and reported in parentheses.
\* p < 0.05, \*\* p < 0.01, \*\*\* p < 0.001.

**Figure 1:**
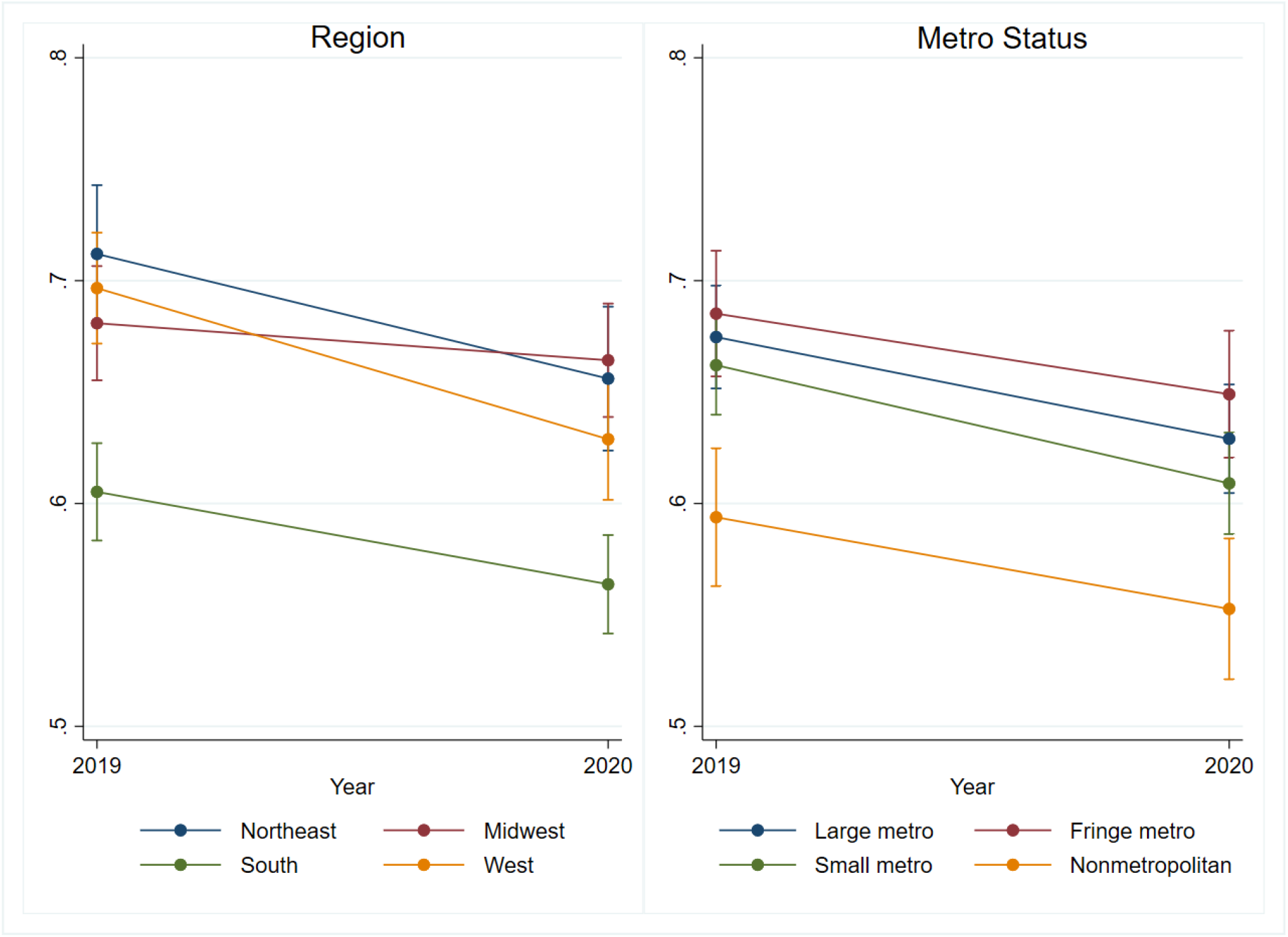
Proportion of Adults Reporting a Dental Visit in the Past Year, by Region and Metro. Figure 1 shows the proportion of adults reporting a a dental visit in 2019 and 2020, by region and Metro Status. Error bars represent 95% confidence interval.

**Figure 2:**
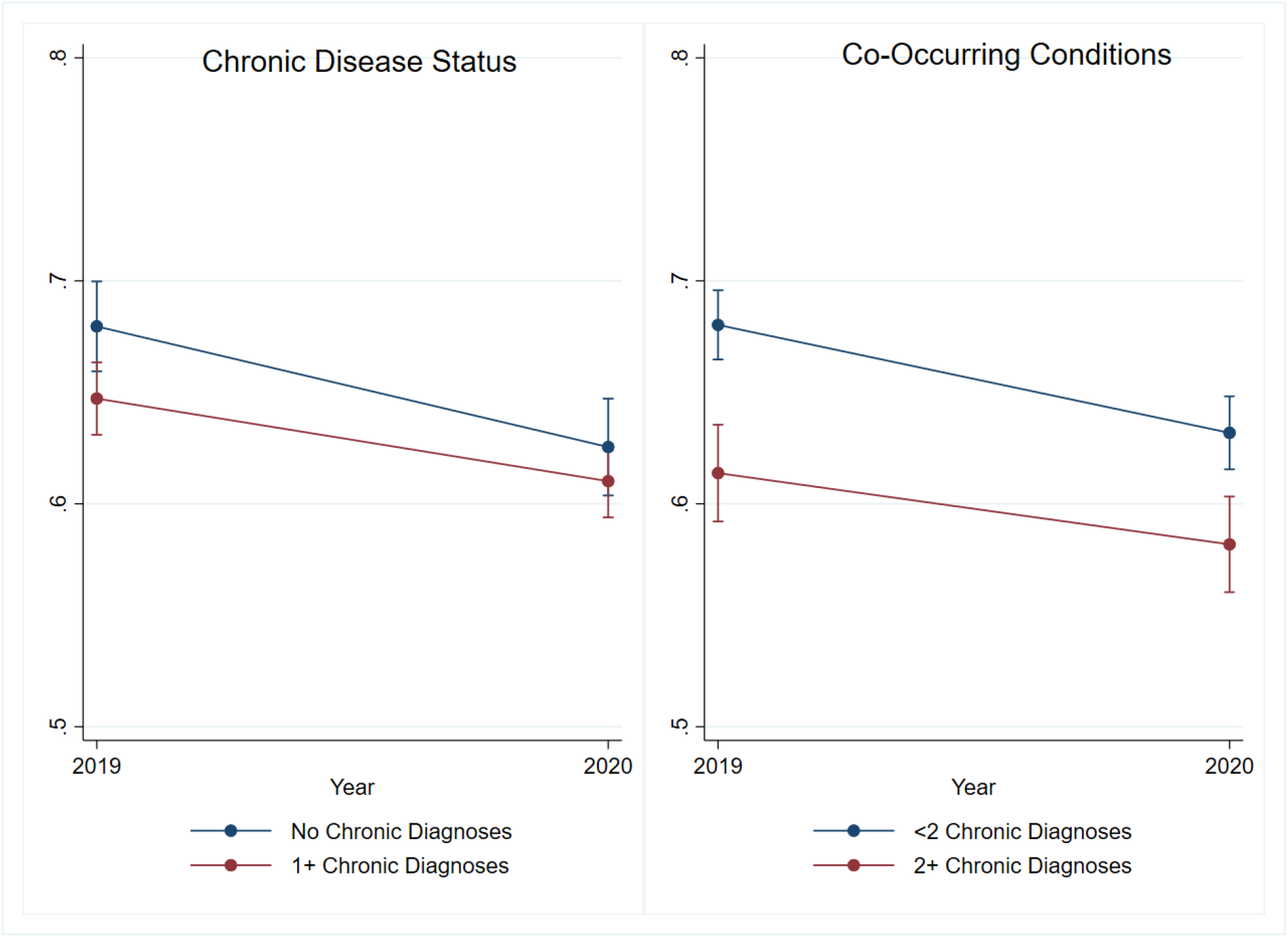
Proportion of Adults Reporting a Dental Visit in the Past Year, by Chronic Disease Status and Co-Occurring Conditions. Figure 2 shows the proportion of adults reporting a a dental visit in 2019 and 2020, by number of chronic disease diagnoses. Error bars represent 95% confidence interval.

**Figure 3:**
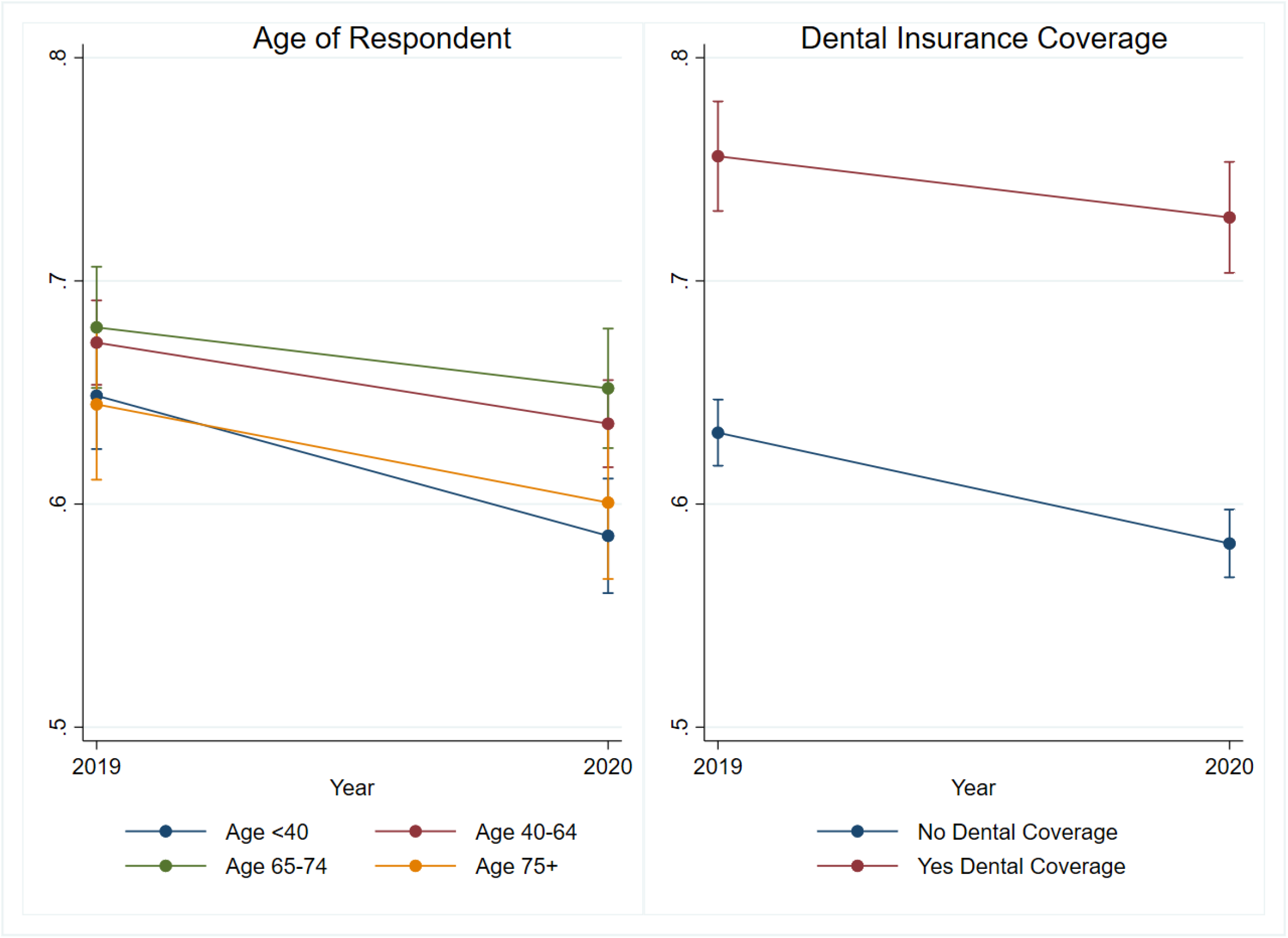
Proportion of Adults Reporting a Dental Visit in the Past Year, by Age and Dental Coverage Status. Figure 3 shows the proportion of adults reporting a a dental visit in 2019 and 2020, by age and dental coverage. Error bars represent 95% confidence interval.

### Changing Dental Service Access Measures

#### Full Sample

First, we find that reported financial barriers to dental care declined by 1.3%-points (p < 0.05), which corresponds to a 6% relative change from 2019. We also find that adults were less likely to report non-financial barriers to care as the proportion of adults reporting an inability to obtain necessary dental care declined by 2.1%-points (p < 0.001). This represents a 12% decline from 2019. Regarding the reports of a most recent dental visit, we see clear evidence that adults were less likely in 2020 to visit the dentist in the past year (Est. = -0.046, p < 0 .001). This represents a 7% relative change from 2019. The decline in a dental visit within the past year appears to be offset by an increase in the proportion of adults reporting a dental visit 1-2 years ago. We find no statistically significant or meaningful changes in reports of a most recent dental visit occurring 2 or more years ago. Similarly, we find no change in the proportion of adults reporting to have never visited a dentist. Finally, there appears to be no difference in the proportion of adults reporting private dental coverage in 2019 compared to 2020.

#### By Exposure to the pandemic

When estimating the within-person change of visiting the dentist in the past year, we find significant heterogeneity by region. As predicted, we observe the largest declines for adults living in the Northeast (Est. = - 0.061, p < 0.001) and West (Est. = -0.069, p < 0.001) regions. Both of these estimates are significantly different than the estimated change for adults in the Midwest (Est. = -0.018, se = 0.013). We also identified a significant difference between the estimate for adults in the West compared to the estimate for adults in the South (Est. = -0.039, p < 0.001).

Contrary to our prediction, we do not find any evidence to suggest that dental access measures and visit patterns differentially changed in 2020 by metro status. The estimated decline in the proportion of adults visiting the dentist in the past year was largest in small metro areas (Est. = -0.054, p < 0.001) and smallest in fringe metro areas (Est. = -.039, p < 0.05). We fail to reject the null hypotheses that the estimates are significantly different across metro status. Regarding other access measures, the only significant difference by metro status was found for the decline in the proportion of adults delaying dental care due to cost in large metro (Est. = -0.001, se = 0.011) compared to non-metro areas (Est. = -0.029, p < 0.05).

#### By Risk of Adverse COVID-19 Outcomes

We predicted that higher risk of adverse COVID-19 outcomes (i.e., older age, more chronic conditions) would be associated with higher declines in dental visits in 2020. However, we find evidence contrary to this prediction. In 2020, adults without a chronic diagnosis were 6.1%-points less likely (p < 0.001) to visit the dentist in the past year, compared to just a 4.0%-point decline (p < 0.001) in adults with at least one chronic diagnosis.

Similarly, the change in proportion of adults visiting the dentist in the past year was larger in adults with one or fewer chronic diagnoses (Est. = -0.052, p < 0.001) than the estimate for adults with two or more chronic diagnoses (Est. = -0.032, p < 0.01). For both sets of estimates, we reject the null hypothesis that the higher risk group had larger declines in dental service visits. However, we do find significantly larger declines in other access measures for the two higher risk groups.

Across the four age groups (18-44, 45-64, 65-75, 75-84), we find little evidence that older age was associated with larger declines in dental visits. Contrary to our hypothesis, the largest decline was estimated for the youngest group of adults age 18-44 (Est. = 0.061, p < 0.001) and the smallest decline was estimated for adults age 45-64 (Est. = -0.037, p < 0.001). Only when comparing the estimates for these two groups did find significantly different estimates. Regarding other access measures, only for the youngest age group did we detect significantly different estimates than changes in other age groups.

#### By Propensity to Visit a Dentist

While overall rates were significantly different between adults with and without dental coverage, we fail to reject the null hypothesis that dental coverage was associated with smaller declines in dental visits in 2020 (Est. No Coverage = -0.050, p < 0.001, se = 0.009; Est. Yes Coverage = -0.042, p < 0.05, se = 0.019). The other two access measures also did not differ by dental insurance status. However, the estimated declines relative to baseline rates were larger in adults without dental coverage (9%) compared to the relative change in adults with dental coverage (5%).

## Discussion

Like most “non-essential” healthcare services in 2020, dental services declined substantially in March and April 2020, resulting in an overall decline compared to 2019 [14-17]. Adding to the prior evidence base, our study contributes an internally valid estimate measuring the within-person changes in dental access measures in 2020 and found significant declines in reports of a dental visit in the past year. However, it is our second aim which may be most interesting to policymakers going forward. With the goal of informing mechanisms for reducing delayed dental treatment, we aimed to identify who was most likely to delay dental care in 2020 and, more importantly, understand why.

We report no evidence to support the claim that dental service visits declined because of greater financial barriers to care in 2020, compared to 2019. In fact, for the full sample and most subgroups, we find a lower proportion of adults reporting delayed dental care because of cost. Now, this conclusion should not be interpreted to infer that the pandemic, or any other event in 2020, addressed financial barriers to dental care. Similarly, we find no evidence that, in 2020 compared to 2019, adults were more likely to report non-financial access barriers or inability to obtain necessary or desired dental care. Again, these results should not be interpreted to indicate that dental capacity to provide services increased in 2020 or that adults were met with fewer administrative and process barriers to accessing dental care. The reported declines in the proportion of adults delaying dental care due to cost and reporting an inability to obtain care are most likely due to the lower propensity and interest in visiting a dentist in 2020: Whether because of social distancing behavior or risk avoidance, adults in 2020 chose to delay dental care because of the COVID-19 pandemic.

Here is where we use the results of who delayed dental care in 2020 to help us understand the “why”. We also use this section to draw attention to the pre and post-COVID disparities in annual dental visits. First, we start with the expectation of visiting a dentist based on the individual’s decision to obtain dental insurance coverage. As stated above, there are no significant differences in dental visit patterns based on dental coverage, and neither was the pandemic associated with changing rates of dental coverage enrollment. However, the overall differences in utilization are stark. Adults with dental coverage were over 10%-points more likely to visit the dentist in the past year compared to adults without coverage. Continued expansion of affordable dental coverage, whether through Medicaid Expansion paired with Medicaid dental coverage, subsidized dental insurance markets, or adults Medicare dental coverage should be explored for their potential efficacy in reducing dental care delays in 2021 and beyond.

It appears that the declines in dental services were not necessarily mediated by individual’s aversion to adverse COVID-19 outcomes based on predisposing risk factors. Our predictions that age and chronic disease status would be associated with higher declines in dental visits were incorrect. In fact, we observed the opposite: that higher declines in dental visits were associated with young ages and adults without chronic diagnoses. We leave the potential explanations for these findings to the reader and future researchers.

Conversely, there does appear to be evidence that exposure to the pandemic was associated with larger declines in dental visits. Unfortunately, given data limitations, we cannot infer whether this association was due to higher case rates, closures, lockdown policies, or social distancing norms. All we can say with confidence is that, n average, case rates, lockdown mandates and social distancing norms were higher in Northeast and West regions compared to Midwest and South [18]. Declines in dental services were higher in the NE/W regions. In fact, we do not find any evidence that dental visits declined in 2020, compared to 2019, for adults in the Midwest. The larger declines in the NE/W compared to the MW may have, in fact, mitigated or even removed completely regional disparities in annual dental visits. However, regional disparities could return or even intensify if the post-COVID rebound in dental services lags in Midwest or South (which has the lowest rates of adults reporting a dental visit in the past year for both 2019 and 2020). Similarly, while there were no differences in the decline by metro status, if the post-COVID dental service rebound lags in nonmetro areas, we would expect rural-urban disparities in dental service and oral health to promulgate over the coming years.

Delayed and forgone dental care can lead to gum disease, tooth loss, and mouth pain, however policies which increase access to dental care can reduce unmet needs and improve oral health [22]. Knowing that dental services declined, for the most part evenly across the population, should motivate policymakers and advocates of multiple constituencies to enact policies and implement or fund programs which promote returning to the dentist. Failing to do so could exacerbate oral health disparities between regions and populations. Finally, for the research community, along with continued monitoring of the dental service and oral health status of adults as we enter the third year of life after COVID, our study should motivate future research using the pandemic’s “shock” to the dental healthcare system to rigorously examine how delays of care impact, not only oral health status, but general health and wellbeing indicators. Such evidence could continue to highlight the value and limits of dental treatment as a tool for improving quality of life and ending health disparities.

### Limitations

This study is not without its limitations. By including individual-level fixed-effects, this within-person design accounts for time-invariant, unobservable differences across individuals which may be influencing dental service utilization. Unfortunately, we cannot rule out the possibility that the global shock induced by the COVID-19 pandemic systematically altered individual’s propensity to visit the dentist, differently by unobservable and unmeasured factors. Because of this possibility, we cannot say with certainty that the changes we estimate in dental service visits were caused by direct effects of the COVID-19 pandemic (i.e., closures, lockdowns, social distancing) or indirect effects (i.e., the pandemic lowered mental health status differentially across the population, which had downstream impacts on subgroups of adults’ dental service utilization patterns). And, while we are confident that our results suggesting that financial barriers and dental provider capacity were not the primary causes of lower dental visits in 2020, we base those conclusions on self-reported dental access measures which could be internalized differently by respondents in 2019 and 2020. Related to our subgroup analyses, we focused on populations which could inform our understanding of the mechanisms for delaying dental care during 2020 and did not necessarily focus on adults with the lowest oral health status or highest rates of dental care delays. Given the limited sample, we did not necessarily have the power to detect effects of the pandemic on dental service outcomes for socioeconomic minorities and vulnerable populations. Future research investigating the long-term trends in dental services should prioritize such groups, most importantly to identify (and respond to) disproportionate rebounds in dental service patterns after the initial pandemic year.

## Conclusions

Using population-based data and a within-person study design, we estimate that adults in 2020 were 4.6%-points less likely to visit the dentist compared to 2019. This change represents a 7% relative change from 2019. The reduced probability of visiting the dentist in 2020 does not appear to be purely driven by individual choices for delaying dental care, as we found no evidence that individuals were more likely to report increased financial barriers to dental care or increased inability to access dental services in 2020. We do find some evidence that the decision to delay dental care in 2020 were mediated by exposure to the pandemic, as larger changes were found in regions with greater exposure to the pandemic’s effect (NE, W) compared to regions with less exposure (MW, S). Contrary to the predictions based on our conceptual model, we did not find evidence that delaying dental services in 2020 was increasingly associated with more chronic diseases, older age, and lack of dental insurance coverage. Whether and how soon dental service patterns experience a post-COVID rebound remains to be seen. The long-term effects of the COVID-19 pandemic on disparities in delayed dental care and oral health warrants continued monitoring as policymakers aim to mitigate the pandemic’s negative consequences on oral health and oral health equity.

## Supporting information

Replication Files and Data Dictionary

## Data Availability

Data dictionary, IPUMS creation file, and code to clean and analyze data uploaded. Data available for free upon registration with IPUMS-NHIS.

https://nhis.ipums.org/nhis/

