## Supplemental Tables 1-3 for "Estimating the within-person change in dental service access measures during the COVID-19 Pandemic"

**Supplemental Files**

**Supplemental Table 1: Sample Characteristics**

| | Proportion | (se) |
|---|---|---|
| *Northeast* | 0.178 | (0.005) |
| *Midwest* | 0.210 | (0.005) |
| *South* | 0.377 | (0.007) |
| *West* | 0.235 | (0.006) |
| *Large Metro* | 0.316 | (0.006) |
| *Fringe Metro* | 0.236 | (0.006) |
| *Small Metro* | 0.305 | (0.006) |
| *Nonmetro* | 0.143 | (0.004) |
| *No Chronic Diagnoses* | 0.449 | (0.007) |
| *1+ Chronic Diagnoses* | 0.551 | (0.007) |
| *No Co-Occurring Diagnoses* | 0.722 | (0.006) |
| *2+ Chronic Diagnoses* | 0.278 | (0.006) |
| *Age < 40 Years* | 0.383 | (0.007) |
| *Age 40-64 Years* | 0.406 | (0.006) |
| *Age 65-74 Years* | 0.125 | (0.004) |
| *Age 75+ Years* | 0.086 | (0.003) |

*Supplemental table 1 reports the summary statistics (characteristics) of the analytical sample. Each subgroup category contains mutually exclusive groups. (N = 10,415 individuals).*

**Supplemental Table 2: Baseline rates of dental service outcomes (2019)**

| | Delayed Dental Care Due to Cost | | Unable to Receive Dental Care | | Last Visit <1 Year | | Has Dental Coverage | |
|---|---|---|---|---|---|---|---|---|
| | **Est.** | **(se)** | **Est.** | **(se)** | **Est.** | **(se)** | **Est.** | **(se)** |
| *Full Sample* | 0.220 | (0.006) | 0.180 | (0.005) | 0.662 | (0.007) | 0.241 | (0.006) |
| *Northeast* | 0.179 | (0.014) | 0.158 | (0.014) | 0.712 | (0.016) | 0.221 | (0.014) |
| *Midwest* | 0.183 | (0.011) | 0.139 | (0.009) | 0.681 | (0.013) | 0.293 | (0.013) |
| *South* | 0.262 | (0.010) | 0.221 | (0.010) | 0.605 | (0.011) | 0.243 | (0.010) |
| *West* | 0.218 | (0.012) | 0.169 | (0.011) | 0.697 | (0.013) | 0.205 | (0.011) |
| *Large Metro* | 0.217 | (0.011) | 0.186 | (0.010) | 0.675 | (0.012) | 0.254 | (0.011) |
| *Fringe Metro* | 0.196 | (0.013) | 0.155 | (0.011) | 0.685 | (0.014) | 0.259 | (0.013) |
| *Small Metro* | 0.234 | (0.010) | 0.186 | (0.010) | 0.662 | (0.011) | 0.232 | (0.010) |
| *Nonmetro* | 0.240 | (0.014) | 0.197 | (0.013) | 0.594 | (0.016) | 0.199 | (0.013) |
| *No Chronic Diagnoses* | 0.199 | (0.009) | 0.157 | (0.008) | 0.680 | (0.010) | 0.277 | (0.010) |
| *1+ Chronic Diagnoses* | 0.238 | (0.008) | 0.199 | (0.007) | 0.647 | (0.008) | 0.211 | (0.007) |
| *No Co-Occurring Diagnoses* | 0.205 | (0.007) | 0.162 | (0.006) | 0.680 | (0.008) | 0.264 | (0.007) |
| *2+ Chronic Diagnoses* | 0.260 | (0.011) | 0.227 | (0.010) | 0.614 | (0.011) | 0.182 | (0.008) |
| *Age < 40 Years* | 0.224 | (0.011) | 0.178 | (0.010) | 0.649 | (0.012) | 0.288 | (0.012) |
| *Age 40-64 Years* | 0.243 | (0.009) | 0.204 | (0.008) | 0.672 | (0.010) | 0.257 | (0.009) |
| *Age 65-74 Years* | 0.194 | (0.012) | 0.151 | (0.011) | 0.679 | (0.014) | 0.134 | (0.009) |
| *Age 75+ Years* | 0.134 | (0.014) | 0.120 | (0.015) | 0.645 | (0.017) | 0.109 | (0.010) |

Supplemental Table 2 reports the baseline rates (2019) for each primary outcome variable, for the full sample and all subgroups. The proportion is reported, along with the standard error (in parentheses).

**Supplemental Table 3: Sensitivity Checks**

| | Delayed Dental Care Due to Cost | | Unable to Receive Dental Care | | Last Visit <1 Year | |
|---|---|---|---|---|---|---|
| | **Estimate** | **(se)** | **Estimate** | **(se)** | **Estimate** | **(s)** |
| *Primary* | -0.013* | (0.006) | -0.021*** | (0.006) | -0.046*** | (0.007) |
| *Alt. 1* | -0.018*** | (0.004) | -0.021*** | (0.004) | -0.041*** | (0.004) |
| *Alt. 2* | -0.012** | (0.004) | -0.016*** | (0.004) | -0.044*** | (0.005) |

Supplemental Table 3 reports the results of the sensitivity checks, for the full sample of adults. The primary model estimates the within-person change in dental service outcomes computing a weighted linear probability model with individual-level fixed effects. The Alt. 1 model removes the weights from the primary analysis. The Alt. 2 model removes the weights and estimates the between person change by computing a linear probability model with individual-level random effects and control variables (age, gender, marital status, race/ethnicity, education attainment, employment status, insurance status, sick leave status, household income, and federal poverty level). We fail to reject the null hypothesis that the results of these alternative specifications are significantly different than the results of our primary specification. * $p < 0.05$, ** $p < 0.01$, *** $p < 0.001$.
