## Supplementary material for "Estimating the within-person change in dental service access measures during the COVID-19 Pandemic": Replication Files and Data Dictionary: nhis_00002.do

```
* NOTE: You need to set the Stata working directory to the path
* where the data file is located.
cd "C:\Users\jsemprini\OneDrive - University of Iowa\4-Misc Projects\d-NHIS-covid\nhis_raw"
set more off

clear
quietly infix                    ///
  int     year          1-4      ///
  long    serial        5-10     ///
  byte    quarter       11-11    ///
  int     strata        12-15    ///
  int     psu           16-18    ///
  str     nhishid       19-32    ///
  byte    intervwqtr    33-33    ///
  byte    region        34-35    ///
  byte    urbrrl        36-36    ///
  byte    pernum        37-38    ///
  str     nhispid       39-54    ///
  str     nhispidprvyr  55-70    ///
  str     hhx           71-77    ///
  double  sampweight    78-89    ///
  double  longweight    90-100   ///
  double  partweight    101-111  ///
  byte    astatflg      112-112  ///
  byte    cstatflg      113-113  ///
  int     age           114-116  ///
  byte    sex           117-117  ///
  byte    marstat       118-119  ///
  byte    marst         120-121  ///
  int     racea         122-124  ///
  byte    hispeth       125-126  ///
  byte    yrsinus       127-127  ///
  byte    usborn        128-129  ///
  byte    citizen       130-130  ///
  int     racenew       131-133  ///
  int     educ          134-136  ///
  byte    spouseduc     137-138  ///
  int     empstat       139-141  ///
  byte    hourswrk      142-143  ///
  byte    paidsick      144-144  ///
  byte    emphi         145-145  ///
  byte    empft         146-146  ///
  byte    emplast       147-147  ///
  byte    incfam07on    148-149  ///
  long    famtotinc     150-155  ///
  byte    qtcincfam     156-156  ///
  byte    impincfam     157-157  ///
  double  cpi2009       158-161  ///
  byte    poverty       162-163  ///
  double  bmicalc       164-167  ///
  byte    bmicat        168-168  ///
  byte    pregnantnow   169-169  ///
  byte    livebirth     170-170  ///
  byte    delaydt       171-171  ///
  byte    unmtnddt      172-172  ///
  byte    hinotcove     173-173  ///
  byte    hiprivatee    174-174  ///
  byte    hichipe       175-176  ///
  byte    himilite      177-178  ///
  byte    hiothgove     179-180  ///
  byte    histatee      181-182  ///
  byte    himcaide      183-183  ///
  byte    himcaree      184-184  ///
  byte    sdentale      185-185  ///
  byte    hinotcov      186-187  ///
  byte    hiprivate     188-189  ///
  byte    hilast        190-191  ///
  byte    hinotyr       192-192  ///
  byte    hinotyrmo     193-194  ///
  byte    hip1dental    195-195  ///
  byte    opioidapain   196-196  ///
  byte    opioidcpain   197-197  ///
  int     dentint       198-200  ///
  byte    asthmaev      201-201  ///
  byte    asthmastil    202-202  ///
  byte    cancerev      203-203  ///
  byte    cheartdiev    204-204  ///
  byte    cholhighev    205-205  ///
  byte    cholhighyr    206-206  ///
  byte    dementiaev    207-207  ///
  byte    diabeticev    208-208  ///
  byte    heartattev    209-209  ///
  byte    hypertenev    210-210  ///
  byte    hypertenyr    211-211  ///
  byte    strokev       212-212  ///
  byte    copdev        213-213  ///
  byte    painfreq3mo   214-214  ///
  byte    painfeelevl   215-215  ///
  byte    paintooth3m   216-216  ///
  byte    cvddiag       217-217  ///
  byte    cvdtest       218-218  ///
  byte    cvdtestrslt   219-219  ///
  byte    cvdsymp       220-220  ///
  byte    cvdcanothtr   221-221  ///
  byte    cvddlycare    222-222  ///
  byte    cvddngcare    223-223  ///
  using `"nhis_00002.dat"'

replace sampweight   = sampweight   / 1000
replace longweight   = longweight   / 1000
replace partweight   = partweight   / 1000
replace cpi2009      = cpi2009      / 1000
replace bmicalc      = bmicalc      / 10

format sampweight   %12.3f
format longweight   %11.3f
format partweight   %11.3f
format cpi2009      %4.3f
format bmicalc      %4.1f

label var year         `"Survey year"'
label var serial       `"Sequential Serial Number, Household Record"'
label var quarter      `"Sample quarter, household record"'
label var strata       `"Stratum for variance estimation"'
label var psu          `"Primary sampling unit (PSU) for variance estimation"'
label var nhishid      `"NHIS Unique identifier, household"'
label var intervwqtr   `"Calendar quarter of NHIS interview"'
label var region       `"Region of residence"'
label var urbrrl       `"Urban-Rural county classification"'
label var pernum       `"Person number within family/household (from reformatting)"'
label var nhispid      `"NHIS Unique Identifier, person"'
label var nhispidprvyr `"NHIS unique identifier for previous year, person"'
label var hhx          `"Household number (from NHIS)"'
label var sampweight   `"Sample Person Weight"'
label var longweight   `"Sample adult weight, longitudinal sample"'
label var partweight   `"Sample adult weight, partial sample"'
label var astatflg     `"Sample adult flag"'
label var cstatflg     `"Sample child flag"'
label var age          `"Age"'
label var sex          `"Sex"'
label var marstat      `"Legal marital status"'
label var marst        `"Current marital status"'
label var racea        `"Main Racial Background (Pre-1997 Revised OMB Standards), self-reported or interv"'
label var hispeth      `"Hispanic ethnicity"'
label var yrsinus      `"Number of years spent in the U.S."'
label var usborn       `"Born in the United States"'
label var citizen      `"U.S. citizenship"'
label var racenew      `"Self-reported Race (Post-1997 OMB standards)"'
label var educ         `"Educational attainment"'
label var spouseduc    `"Education level of sample adult's spouse"'
label var empstat      `"Employment status in past 1 to 2 weeks"'
label var hourswrk     `"Total hours worked last week or usually"'
label var paidsick     `"Paid sick leave at current job"'
label var emphi        `"Workplace offered health insurance"'
label var empft        `"Usually work 35+ hours per week"'
label var emplast      `"Last time worked for pay"'
label var incfam07on   `"Total combined family income (2007+)"'
label var famtotinc    `"Total family income, last year (top coded)"'
label var qtcincfam    `"Family income top code flag"'
label var impincfam    `"Family income imputation flag"'
label var cpi2009      `"CPI conversion factor"'
label var poverty      `"Ratio of family income to poverty threshold"'
label var bmicalc      `"Body Mass Index, calculated from publicly released height and weight variables"'
label var bmicat       `"Categorical body mass index"'
label var pregnantnow  `"Currently pregnant"'
label var livebirth    `"Had live birth in the past 12 months"'
label var delaydt      `"Delayed getting dental care, past 12 months"'
label var unmtnddt     `"Unable to get needed dental care"'
label var hinotcove    `"Health Insurance coverage status"'
label var hiprivatee   `"Covered by private health insurance: Recode"'
label var hichipe      `"Covered by Children's Health Insurance Program: Recode"'
label var himilite     `"Covered by military health insurance: Recode"'
label var hiothgove    `"Covered by other government program: Recode"'
label var histatee     `"Covered by other state-sponsored health plan: Recode"'
label var himcaide     `"Covered by Medicaid: Recode"'
label var himcaree     `"Covered by Medicare: Recode"'
label var sdentale     `"Single service plan for dental care"'
label var hinotcov     `"Has no health insurance (excluding single service plans)"'
label var hiprivate    `"Has any private health insurance"'
label var hilast       `"Time since had health coverage (no coverage now)"'
label var hinotyr      `"Had no health coverage at some point during past 12 months"'
label var hinotyrmo    `"Months without health coverage, past 12 months"'
label var hip1dental   `"Plan 1: Has dental coverage"'
label var opioidapain  `"Used prescription opioids for acute pain during the past 3 months"'
label var opioidcpain  `"Used prescription opioids for chronic pain during the past 3 months"'
label var dentint      `"Interval since last dental visit"'
label var asthmaev     `"Ever told had asthma"'
label var asthmastil   `"Still have asthma"'
label var cancerev     `"Ever told had cancer"'
label var cheartdiev   `"Ever told had coronary heart disease"'
label var cholhighev   `"Ever told had high cholesterol"'
label var cholhighyr   `"Had high cholesterol, past 12 months"'
label var dementiaev   `"Ever told you had Dementia"'
label var diabeticev   `"Ever told had diabetes"'
label var heartattev   `"Ever told had heart attack"'
label var hypertenev   `"Ever told had hypertension"'
label var hypertenyr   `"Had hypertension, past 12 months"'
label var strokev      `"Ever told had a stroke"'
label var copdev       `"Ever been told you had COPD"'
label var painfreq3mo  `"Frequency of pain in last 3 months"'
label var painfeelevl  `"Level of pain, last time had pain"'
label var paintooth3m  `"How much jaw or tooth pain, past 3 months"'
label var cvddiag      `"Ever told had or likely had coronavirus"'
label var cvdtest      `"Ever been tested for coronavirus"'
label var cvdtestrslt  `"Had coronavirus, according to test"'
label var cvdsymp      `"Severity of coronavirus symptoms at their worst"'
label var cvdcanothtr  `"Needed other medical care for cancer since start of coronavirus pandemic"'
label var cvddlycare   `"Delayed getting medical care because of the coronavirus pandemic"'
label var cvddngcare   `"Did not get needed medical care other than for coronavirus because of the corona"'

label define quarter_lbl 1 `"Quarter 1"'
label define quarter_lbl 2 `"Quarter 2"', add
label define quarter_lbl 3 `"Quarter 3"', add
label define quarter_lbl 4 `"Quarter 4"', add
label define quarter_lbl 9 `"Unknown"', add
label values quarter quarter_lbl

label define intervwqtr_lbl 1 `"Quarter 1"'
label define intervwqtr_lbl 2 `"Quarter 2"', add
label define intervwqtr_lbl 3 `"Quarter 3"', add
label define intervwqtr_lbl 4 `"Quarter 4"', add
label values intervwqtr intervwqtr_lbl

label define region_lbl 01 `"Northeast"'
label define region_lbl 02 `"North Central/Midwest"', add
label define region_lbl 03 `"South"', add
label define region_lbl 04 `"West"', add
label define region_lbl 08 `"NO DATA IN ROUND"', add
label define region_lbl 09 `"Unknown"', add
label values region region_lbl

label define urbrrl_lbl 1 `"Large central metro"'
label define urbrrl_lbl 2 `"Large fringe metro"', add
label define urbrrl_lbl 3 `"Medium and small metro"', add
label define urbrrl_lbl 4 `"Nonmetropolitan"', add
label values urbrrl urbrrl_lbl

label define astatflg_lbl 0 `"NIU"'
label define astatflg_lbl 1 `"Sample adult, has record"', add
label define astatflg_lbl 2 `"Sample adult, no record"', add
label define astatflg_lbl 3 `"Not selected as sample adult"', add
label define astatflg_lbl 4 `"No one selected as sample adult"', add
label define astatflg_lbl 5 `"Armed forces member"', add
label define astatflg_lbl 6 `"AF member, selected as sample adult"', add
label values astatflg astatflg_lbl

label define cstatflg_lbl 0 `"NIU"'
label define cstatflg_lbl 1 `"Sample child-has record"', add
label define cstatflg_lbl 2 `"Sample child-no record"', add
label define cstatflg_lbl 3 `"Not selected as sample child"', add
label define cstatflg_lbl 4 `"No one selected as sample child"', add
label define cstatflg_lbl 5 `"Emancipated minor"', add
label values cstatflg cstatflg_lbl

label define sex_lbl 1 `"Male"'
label define sex_lbl 2 `"Female"', add
label define sex_lbl 7 `"Unknown-refused"', add
label define sex_lbl 8 `"Unknown-not ascertained"', add
label define sex_lbl 9 `"Unknown-don't know"', add
label values sex sex_lbl

label define marstat_lbl 00 `"NIU"'
label define marstat_lbl 10 `"Married"', add
label define marstat_lbl 11 `"Married - Spouse present"', add
label define marstat_lbl 12 `"Married - Spouse not in household"', add
label define marstat_lbl 13 `"Married - Spouse in household unknown"', add
label define marstat_lbl 20 `"Widowed"', add
label define marstat_lbl 30 `"Divorced"', add
label define marstat_lbl 40 `"Separated"', add
label define marstat_lbl 50 `"Never married"', add
label define marstat_lbl 99 `"Unknown marital status"', add
label values marstat marstat_lbl

label define marst_lbl 00 `"NIU"'
label define marst_lbl 10 `"Married"', add
label define marst_lbl 11 `"Married - Spouse present"', add
label define marst_lbl 12 `"Married - Spouse not in household"', add
label define marst_lbl 13 `"Married - Spouse in household unknown"', add
label define marst_lbl 20 `"Widowed"', add
label define marst_lbl 30 `"Divorced"', add
label define marst_lbl 40 `"Separated"', add
label define marst_lbl 50 `"Never married"', add
label define marst_lbl 99 `"Unknown marital status"', add
label values marst marst_lbl

label define racea_lbl 100 `"White"'
label define racea_lbl 200 `"Black/African-American"', add
label define racea_lbl 300 `"Aleut, Alaskan Native, or American Indian"', add
label define racea_lbl 310 `"Alaskan Native or American Indian"', add
label define racea_lbl 320 `"Alaskan Native/Eskimo"', add
label define racea_lbl 330 `"Aleut"', add
label define racea_lbl 340 `"American Indian"', add
label define racea_lbl 350 `"American Indian or Alaskan Native and any other group"', add
label define racea_lbl 400 `"Asian or Pacific Islander"', add
label define racea_lbl 410 `"Asian"', add
label define racea_lbl 411 `"Chinese"', add
label define racea_lbl 412 `"Filipino"', add
label define racea_lbl 413 `"Korean"', add
label define racea_lbl 414 `"Vietnamese"', add
label define racea_lbl 415 `"Japanese"', add
label define racea_lbl 416 `"Asian Indian"', add
label define racea_lbl 420 `"Pacific Islander"', add
label define racea_lbl 421 `"Hawaiian"', add
label define racea_lbl 422 `"Samoan"', add
label define racea_lbl 423 `"Guamanian"', add
label define racea_lbl 430 `"Other Asian or Pacific Islander"', add
label define racea_lbl 431 `"Other Asian or Pacific Islander (1992-1995)"', add
label define racea_lbl 432 `"Other Asian or Pacific Islander (1996)"', add
label define racea_lbl 433 `"Other Asian or Pacific Islander (1997-1998)"', add
label define racea_lbl 434 `"Other Asian (1999 forward)"', add
label define racea_lbl 500 `"Other Race"', add
label define racea_lbl 510 `"Other Race (1963-1977)"', add
label define racea_lbl 520 `"Other Race (1978)"', add
label define racea_lbl 530 `"Other Race (1979-1991)"', add
label define racea_lbl 540 `"Other Race (1992-1995)"', add
label define racea_lbl 550 `"Other Race (1996)"', add
label define racea_lbl 560 `"Other Race (1997-1998)"', add
label define racea_lbl 570 `"Other Race (1999-2002)"', add
label define racea_lbl 580 `"Primary Race not releasable"', add
label define racea_lbl 590 `"Other Single and Multiple Races"', add
label define racea_lbl 600 `"Multiple Race, No Primary Race Selected"', add
label define racea_lbl 610 `"Multiple Race, including Asian, excluding Black and White"', add
label define racea_lbl 611 `"Multiple Race, including Asian and Black, excluding White"', add
label define racea_lbl 612 `"Multiple Race, including Asian and White, excluding Black"', add
label define racea_lbl 613 `"Multiple Race, including Black, excluding Asian and White"', add
label define racea_lbl 614 `"Multiple Race, including Black and White, excluding Asian"', add
label define racea_lbl 615 `"Multiple Race, including White, excluding Asian and Black"', add
label define racea_lbl 616 `"Multiple Race, including Asian, White, and Black"', add
label define racea_lbl 617 `"Multiple Race, excluding Asian, White, and Black"', add
label define racea_lbl 900 `"Unknown"', add
label define racea_lbl 970 `"Unknown-refused"', add
label define racea_lbl 980 `"Unknown-not ascertained"', add
label define racea_lbl 990 `"Unknown (1997 forward: Don't know)"', add
label values racea racea_lbl

label define hispeth_lbl 10 `"Not Hispanic/Spanish origin"'
label define hispeth_lbl 20 `"Mexican"', add
label define hispeth_lbl 21 `"Mexican-Mexicano"', add
label define hispeth_lbl 22 `"Mexicano"', add
label define hispeth_lbl 23 `"Mexican-American"', add
label define hispeth_lbl 24 `"Chicano"', add
label define hispeth_lbl 30 `"Puerto Rican"', add
label define hispeth_lbl 40 `"Cuban/Cuban American"', add
label define hispeth_lbl 50 `"Dominican (Republic)"', add
label define hispeth_lbl 60 `"Other Hispanic"', add
label define hispeth_lbl 61 `"Central or South American"', add
label define hispeth_lbl 62 `"Other Latin American, type not specified"', add
label define hispeth_lbl 63 `"Other Spanish"', add
label define hispeth_lbl 64 `"Hispanic/Latino/Spanish, non-specific type"', add
label define hispeth_lbl 65 `"Hispanic/Latino/Spanish, type refused"', add
label define hispeth_lbl 66 `"Hispanic/Latino/Spanish, type not ascertained"', add
label define hispeth_lbl 67 `"Hispanic/Spanish, type don't know"', add
label define hispeth_lbl 70 `"Multiple Hispanic"', add
label define hispeth_lbl 90 `"Unknown"', add
label define hispeth_lbl 91 `"Unknown if Hispanic/Spanish origin"', add
label define hispeth_lbl 92 `"Two origins, unknown which is the main"', add
label define hispeth_lbl 93 `"Origin unknown, refused or not reported"', add
label define hispeth_lbl 99 `"NIU"', add
label values hispeth hispeth_lbl

label define yrsinus_lbl 0 `"NIU"'
label define yrsinus_lbl 1 `"Less than 1 year"', add
label define yrsinus_lbl 2 `"1 year to less than 5 years."', add
label define yrsinus_lbl 3 `"5 years to less than 10 years"', add
label define yrsinus_lbl 4 `"10 years to less than 15 years"', add
label define yrsinus_lbl 5 `"15 years or more"', add
label define yrsinus_lbl 7 `"Refused"', add
label define yrsinus_lbl 8 `"Don't Know"', add
label define yrsinus_lbl 9 `"Not Ascertained"', add
label values yrsinus yrsinus_lbl

label define usborn_lbl 10 `"No"'
label define usborn_lbl 11 `"No, born in U.S. territory"', add
label define usborn_lbl 12 `"No, born outside U.S. and U.S. territories"', add
label define usborn_lbl 20 `"Yes, born in U.S."', add
label define usborn_lbl 97 `"Unknown-refused"', add
label define usborn_lbl 98 `"Unknown-not ascertained"', add
label define usborn_lbl 99 `"Unknown-don't know"', add
label define usborn_lbl 96 `"Inapplicable"', add
label values usborn usborn_lbl

label define citizen_lbl 1 `"No, not U.S. citizen"'
label define citizen_lbl 2 `"Yes, U.S. citizen"', add
label define citizen_lbl 7 `"Unknown--refused"', add
label define citizen_lbl 8 `"Unknown--not ascertained"', add
label define citizen_lbl 9 `"Unknown--don't know"', add
label values citizen citizen_lbl

label define racenew_lbl 100 `"White"'
label define racenew_lbl 200 `"Black/African American"', add
label define racenew_lbl 300 `"American Indian/Alaskan Native"', add
label define racenew_lbl 400 `"Asian"', add
label define racenew_lbl 500 `"Other Race and Multiple Race"', add
label define racenew_lbl 510 `"Other Race"', add
label define racenew_lbl 519 `"Race Group Not Releasable"', add
label define racenew_lbl 520 `"Multiple Race"', add
label define racenew_lbl 521 `"Muliple Race, including American Indian/Alaskan Native"', add
label define racenew_lbl 997 `"Unknown-Refused"', add
label define racenew_lbl 998 `"Unknown-Not ascertained"', add
label define racenew_lbl 999 `"Unknown-Don't Know"', add
label values racenew racenew_lbl

label define educ_lbl 000 `"NIU"'
label define educ_lbl 100 `"Grade 12 or less, no high school diploma or equivalent"', add
label define educ_lbl 101 `"Grade 8 or less (no further detail)"', add
label define educ_lbl 102 `"Never attended/kindergarten only"', add
label define educ_lbl 103 `"Grades 1-11 (no further detail)"', add
label define educ_lbl 104 `"Grade 1"', add
label define educ_lbl 105 `"Grade 2"', add
label define educ_lbl 106 `"Grade 3"', add
label define educ_lbl 107 `"Grade 4"', add
label define educ_lbl 108 `"Grade 5"', add
label define educ_lbl 109 `"Grade 6"', add
label define educ_lbl 110 `"Grade 7"', add
label define educ_lbl 111 `"Grade 8"', add
label define educ_lbl 112 `"Grade 9-12, no diploma (no further detail)"', add
label define educ_lbl 113 `"Grade 9"', add
label define educ_lbl 114 `"Grade 10"', add
label define educ_lbl 115 `"Grade 11"', add
label define educ_lbl 116 `"12th grade, no diploma"', add
label define educ_lbl 200 `"High school diploma or GED"', add
label define educ_lbl 201 `"High school graduate"', add
label define educ_lbl 202 `"GED or equivalent"', add
label define educ_lbl 300 `"Some college, no 4yr degree"', add
label define educ_lbl 301 `"Some college, no degree"', add
label define educ_lbl 302 `"AA degree: technical/vocational/occupational"', add
label define educ_lbl 303 `"AA degree: academic program"', add
label define educ_lbl 400 `"Bachelor's degree (BA,AB,BS,BBA)"', add
label define educ_lbl 500 `"Master's, Professional, or Doctoral Degree"', add
label define educ_lbl 501 `"Master's degree (MA,MS,Med,MBA)"', add
label define educ_lbl 502 `"Professional (MD,DDS,DVM,JD)"', add
label define educ_lbl 503 `"Doctoral degree (PhD, EdD)"', add
label define educ_lbl 504 `"Other degree"', add
label define educ_lbl 996 `"No degree, years of education unknown"', add
label define educ_lbl 997 `"Unknown--refused"', add
label define educ_lbl 998 `"Unknown--not ascertained"', add
label define educ_lbl 999 `"Unknown--don't know"', add
label values educ educ_lbl

label define spouseduc_lbl 00 `"NIU"'
label define spouseduc_lbl 01 `"Grade 0-11"', add
label define spouseduc_lbl 02 `"12th grade, no diploma"', add
label define spouseduc_lbl 03 `"GED or equivalent"', add
label define spouseduc_lbl 04 `"High school graduate"', add
label define spouseduc_lbl 05 `"Some college, no degree"', add
label define spouseduc_lbl 06 `"Associate degree (occupational, technical, or vocational program)"', add
label define spouseduc_lbl 07 `"Associate degree (academic program)"', add
label define spouseduc_lbl 08 `"Bachelor's degree"', add
label define spouseduc_lbl 09 `"Master's degree"', add
label define spouseduc_lbl 10 `"Professional school degree"', add
label define spouseduc_lbl 11 `"Doctoral degree"', add
label define spouseduc_lbl 97 `"Unknown - refused"', add
label define spouseduc_lbl 98 `"Unknown - not ascertained"', add
label define spouseduc_lbl 99 `"Unknown - don't know"', add
label values spouseduc spouseduc_lbl

label define empstat_lbl 000 `"NIU"'
label define empstat_lbl 100 `"Employed"', add
label define empstat_lbl 110 `"Working"', add
label define empstat_lbl 111 `"Working for pay at job/business"', add
label define empstat_lbl 112 `"Working, w/out pay, at job/business"', add
label define empstat_lbl 120 `"With job, but not at work"', add
label define empstat_lbl 121 `"With job, not at work: not laid-off, not looking"', add
label define empstat_lbl 122 `"With job, not at work: looking"', add
label define empstat_lbl 200 `"Not employed"', add
label define empstat_lbl 210 `"Unemployed"', add
label define empstat_lbl 211 `"Unemployed: On layoff"', add
label define empstat_lbl 212 `"Unemployed: On layoff and looking"', add
label define empstat_lbl 213 `"Unemployed: Unk if looking or laid off"', add
label define empstat_lbl 214 `"Unemployed: Looking or on layoff"', add
label define empstat_lbl 215 `"Unemployed: Have job to return to"', add
label define empstat_lbl 216 `"Unemployed: Had job during the round"', add
label define empstat_lbl 217 `"Unemployed: No job during reference period"', add
label define empstat_lbl 220 `"Not in labor force"', add
label define empstat_lbl 900 `"Unknown-all causes"', add
label define empstat_lbl 997 `"Unknown-refused"', add
label define empstat_lbl 998 `"Unknown-not ascertained"', add
label define empstat_lbl 999 `"Unknown-don't know"', add
label values empstat empstat_lbl

label define hourswrk_lbl 00 `"NIU"'
label define hourswrk_lbl 01 `"1"', add
label define hourswrk_lbl 02 `"2"', add
label define hourswrk_lbl 03 `"3"', add
label define hourswrk_lbl 04 `"4"', add
label define hourswrk_lbl 05 `"5"', add
label define hourswrk_lbl 06 `"6"', add
label define hourswrk_lbl 07 `"7"', add
label define hourswrk_lbl 08 `"8"', add
label define hourswrk_lbl 09 `"9"', add
label define hourswrk_lbl 10 `"10"', add
label define hourswrk_lbl 11 `"11"', add
label define hourswrk_lbl 12 `"12"', add
label define hourswrk_lbl 13 `"13"', add
label define hourswrk_lbl 14 `"14"', add
label define hourswrk_lbl 15 `"15"', add
label define hourswrk_lbl 16 `"16"', add
label define hourswrk_lbl 17 `"17"', add
label define hourswrk_lbl 18 `"18"', add
label define hourswrk_lbl 19 `"19"', add
label define hourswrk_lbl 20 `"20"', add
label define hourswrk_lbl 21 `"21"', add
label define hourswrk_lbl 22 `"22"', add
label define hourswrk_lbl 23 `"23"', add
label define hourswrk_lbl 24 `"24"', add
label define hourswrk_lbl 25 `"25"', add
label define hourswrk_lbl 26 `"26"', add
label define hourswrk_lbl 27 `"27"', add
label define hourswrk_lbl 28 `"28"', add
label define hourswrk_lbl 29 `"29"', add
label define hourswrk_lbl 30 `"30"', add
label define hourswrk_lbl 31 `"31"', add
label define hourswrk_lbl 32 `"32"', add
label define hourswrk_lbl 33 `"33"', add
label define hourswrk_lbl 34 `"34"', add
label define hourswrk_lbl 35 `"35"', add
label define hourswrk_lbl 36 `"36"', add
label define hourswrk_lbl 37 `"37"', add
label define hourswrk_lbl 38 `"38"', add
label define hourswrk_lbl 39 `"39"', add
label define hourswrk_lbl 40 `"40"', add
label define hourswrk_lbl 41 `"41"', add
label define hourswrk_lbl 42 `"42"', add
label define hourswrk_lbl 43 `"43"', add
label define hourswrk_lbl 44 `"44"', add
label define hourswrk_lbl 45 `"45"', add
label define hourswrk_lbl 46 `"46"', add
label define hourswrk_lbl 47 `"47"', add
label define hourswrk_lbl 48 `"48"', add
label define hourswrk_lbl 49 `"49"', add
label define hourswrk_lbl 50 `"50"', add
label define hourswrk_lbl 51 `"51"', add
label define hourswrk_lbl 52 `"52"', add
label define hourswrk_lbl 53 `"53"', add
label define hourswrk_lbl 54 `"54"', add
label define hourswrk_lbl 55 `"55"', add
label define hourswrk_lbl 56 `"56"', add
label define hourswrk_lbl 57 `"57"', add
label define hourswrk_lbl 58 `"58"', add
label define hourswrk_lbl 59 `"59"', add
label define hourswrk_lbl 60 `"60"', add
label define hourswrk_lbl 61 `"61"', add
label define hourswrk_lbl 62 `"62"', add
label define hourswrk_lbl 63 `"63"', add
label define hourswrk_lbl 64 `"64"', add
label define hourswrk_lbl 65 `"65"', add
label define hourswrk_lbl 66 `"66"', add
label define hourswrk_lbl 67 `"67"', add
label define hourswrk_lbl 68 `"68"', add
label define hourswrk_lbl 69 `"69"', add
label define hourswrk_lbl 70 `"70"', add
label define hourswrk_lbl 71 `"71"', add
label define hourswrk_lbl 72 `"72"', add
label define hourswrk_lbl 73 `"73"', add
label define hourswrk_lbl 74 `"74"', add
label define hourswrk_lbl 75 `"75"', add
label define hourswrk_lbl 76 `"76"', add
label define hourswrk_lbl 77 `"77"', add
label define hourswrk_lbl 78 `"78"', add
label define hourswrk_lbl 79 `"79"', add
label define hourswrk_lbl 80 `"80"', add
label define hourswrk_lbl 81 `"81"', add
label define hourswrk_lbl 82 `"82"', add
label define hourswrk_lbl 83 `"83"', add
label define hourswrk_lbl 84 `"84"', add
label define hourswrk_lbl 85 `"85"', add
label define hourswrk_lbl 86 `"86"', add
label define hourswrk_lbl 87 `"87"', add
label define hourswrk_lbl 88 `"88"', add
label define hourswrk_lbl 89 `"89"', add
label define hourswrk_lbl 90 `"90"', add
label define hourswrk_lbl 91 `"91"', add
label define hourswrk_lbl 92 `"92"', add
label define hourswrk_lbl 93 `"93"', add
label define hourswrk_lbl 94 `"94"', add
label define hourswrk_lbl 95 `"95+ hours"', add
label define hourswrk_lbl 97 `"Unknown--refused"', add
label define hourswrk_lbl 98 `"Unknown--not ascertained"', add
label define hourswrk_lbl 99 `"Unknown--don't know"', add
label values hourswrk hourswrk_lbl

label define paidsick_lbl 0 `"NIU"'
label define paidsick_lbl 1 `"No"', add
label define paidsick_lbl 2 `"Yes"', add
label define paidsick_lbl 7 `"Unknown-refused"', add
label define paidsick_lbl 8 `"Unknown-not ascertained"', add
label define paidsick_lbl 9 `"Unknown-don't know"', add
label values paidsick paidsick_lbl

label define emphi_lbl 0 `"NIU"'
label define emphi_lbl 1 `"No"', add
label define emphi_lbl 2 `"Yes"', add
label define emphi_lbl 7 `"Unknown-refused"', add
label define emphi_lbl 8 `"Unknown-not ascertained"', add
label define emphi_lbl 9 `"Unknown-don't know"', add
label values emphi emphi_lbl

label define empft_lbl 0 `"NIU"'
label define empft_lbl 1 `"No"', add
label define empft_lbl 2 `"Yes"', add
label define empft_lbl 7 `"Unknown-refused"', add
label define empft_lbl 8 `"Unknown-not ascertained"', add
label define empft_lbl 9 `"Unknown-don't know"', add
label values empft empft_lbl

label define emplast_lbl 0 `"NIU"'
label define emplast_lbl 1 `"Within the past 12 months"', add
label define emplast_lbl 2 `"1-5 years ago"', add
label define emplast_lbl 3 `"Over 5 years ago"', add
label define emplast_lbl 4 `"Never worked"', add
label define emplast_lbl 7 `"Unknown-refused"', add
label define emplast_lbl 8 `"Unknown-not ascertained"', add
label define emplast_lbl 9 `"Unknown-don't know"', add
label values emplast emplast_lbl

label define incfam07on_lbl 10 `"$0 - $49,999 (no further detail)"'
label define incfam07on_lbl 11 `"$0 - $34,999"', add
label define incfam07on_lbl 12 `"$35,000 - $49,999"', add
label define incfam07on_lbl 20 `"$50,000  and over (no further detail; available 2007-2013)"', add
label define incfam07on_lbl 21 `"$50,000 - $99,999 (no further detail)"', add
label define incfam07on_lbl 22 `"$50,000 - $74,999"', add
label define incfam07on_lbl 23 `"$75,000 - $99,999"', add
label define incfam07on_lbl 24 `"$100,000 and over"', add
label define incfam07on_lbl 96 `"Undefined"', add
label define incfam07on_lbl 99 `"Unknown"', add
label values incfam07on incfam07on_lbl

label define qtcincfam_lbl 1 `"No"'
label define qtcincfam_lbl 2 `"Yes"', add
label values qtcincfam qtcincfam_lbl

label define impincfam_lbl 0 `"Income reported"'
label define impincfam_lbl 1 `"Income imputed; no other information"', add
label define impincfam_lbl 2 `"Income imputed; reported in income brackets"', add
label values impincfam impincfam_lbl

label define poverty_lbl 10 `"Less than 1.0"'
label define poverty_lbl 11 `"Under 0.50"', add
label define poverty_lbl 12 `"0.50 to 0.74"', add
label define poverty_lbl 13 `"0.75 to 0.99"', add
label define poverty_lbl 14 `"Less than 1.0 (no other detail)"', add
label define poverty_lbl 20 `"1.00 to 1.99"', add
label define poverty_lbl 21 `"1.00 to 1.24"', add
label define poverty_lbl 22 `"1.25 to 1.49"', add
label define poverty_lbl 23 `"1.50 to 1.74"', add
label define poverty_lbl 24 `"1.75 to 1.99"', add
label define poverty_lbl 25 `"1.00 - 1.99 (no further detail)"', add
label define poverty_lbl 30 `"2.00 and over"', add
label define poverty_lbl 31 `"2.00 to 2.49"', add
label define poverty_lbl 32 `"2.50 to 2.99"', add
label define poverty_lbl 33 `"3.00 to 3.49"', add
label define poverty_lbl 34 `"3.50 to 3.99"', add
label define poverty_lbl 35 `"4.00 to 4.49"', add
label define poverty_lbl 36 `"4.50 to 4.99"', add
label define poverty_lbl 37 `"5.00 and over"', add
label define poverty_lbl 38 `"2.00 and over (no other detail)"', add
label define poverty_lbl 98 `"Undefinable"', add
label define poverty_lbl 99 `"Unknown"', add
label values poverty poverty_lbl

label define bmicat_lbl 0 `"NIU"'
label define bmicat_lbl 1 `"Underweight (BMI < 18.5)"', add
label define bmicat_lbl 2 `"Normal weight (BMI >= 18.5 & BMI < 25)"', add
label define bmicat_lbl 3 `"Overweight (BMI >= 25  & BMI < 30)"', add
label define bmicat_lbl 4 `"Obese (BMI 30+)"', add
label define bmicat_lbl 9 `"Unknown"', add
label values bmicat bmicat_lbl

label define pregnantnow_lbl 0 `"NIU"'
label define pregnantnow_lbl 1 `"No"', add
label define pregnantnow_lbl 2 `"Yes"', add
label define pregnantnow_lbl 7 `"Unknown--refused"', add
label define pregnantnow_lbl 8 `"Unknown--not ascertained"', add
label define pregnantnow_lbl 9 `"Unknown--don't know"', add
label values pregnantnow pregnantnow_lbl

label define livebirth_lbl 0 `"NIU"'
label define livebirth_lbl 1 `"No"', add
label define livebirth_lbl 2 `"Yes"', add
label define livebirth_lbl 7 `"Unknown - refused"', add
label define livebirth_lbl 8 `"Unknown - not ascertained"', add
label define livebirth_lbl 9 `"Unknown - don't know"', add
label values livebirth livebirth_lbl

label define delaydt_lbl 0 `"Not in universe"'
label define delaydt_lbl 1 `"No"', add
label define delaydt_lbl 2 `"Yes"', add
label define delaydt_lbl 7 `"Unknown-refused"', add
label define delaydt_lbl 8 `"Unknown-not ascertained"', add
label define delaydt_lbl 9 `"Unknown-don't know"', add
label values delaydt delaydt_lbl

label define unmtnddt_lbl 0 `"Not in universe"'
label define unmtnddt_lbl 1 `"No"', add
label define unmtnddt_lbl 2 `"Yes"', add
label define unmtnddt_lbl 7 `"Unknown - refused"', add
label define unmtnddt_lbl 8 `"Unknown - not ascertained"', add
label define unmtnddt_lbl 9 `"Unknown - don't know"', add
label values unmtnddt unmtnddt_lbl

label define hinotcove_lbl 0 `"NIU"'
label define hinotcove_lbl 1 `"No, has coverage"', add
label define hinotcove_lbl 2 `"Yes, has no coverage"', add
label define hinotcove_lbl 7 `"Unknown-refused"', add
label define hinotcove_lbl 8 `"Unknown-not ascertained"', add
label define hinotcove_lbl 9 `"Unknown-don't know"', add
label values hinotcove hinotcove_lbl

label define hiprivatee_lbl 1 `"No"'
label define hiprivatee_lbl 2 `"Yes, information"', add
label define hiprivatee_lbl 3 `"Yes, but no information"', add
label define hiprivatee_lbl 7 `"Unknown-refused"', add
label define hiprivatee_lbl 8 `"Unknown-not ascertained"', add
label define hiprivatee_lbl 9 `"Unknown-don't know"', add
label values hiprivatee hiprivatee_lbl

label define hichipe_lbl 10 `"No"'
label define hichipe_lbl 20 `"Yes"', add
label define hichipe_lbl 21 `"Yes, information"', add
label define hichipe_lbl 22 `"Yes, but no information"', add
label define hichipe_lbl 97 `"Unknown-refused"', add
label define hichipe_lbl 98 `"Unknown-not ascertained"', add
label define hichipe_lbl 99 `"Unknown-don't know"', add
label values hichipe hichipe_lbl

label define himilite_lbl 10 `"No"'
label define himilite_lbl 20 `"Yes"', add
label define himilite_lbl 21 `"Yes, Military/VA only"', add
label define himilite_lbl 22 `"Yes, CHAMPUS/TRICARE/CHAMP-VA only"', add
label define himilite_lbl 23 `"Yes, both Military/VA and CHAMPUS/TRICARE"', add
label define himilite_lbl 24 `"Yes, unknown type"', add
label define himilite_lbl 25 `"Yes, information"', add
label define himilite_lbl 26 `"Yes, no information"', add
label define himilite_lbl 97 `"Unknown-refused"', add
label define himilite_lbl 98 `"Unknown-not ascertained"', add
label define himilite_lbl 99 `"Unknown-don't know"', add
label values himilite himilite_lbl

label define hiothgove_lbl 10 `"No"'
label define hiothgove_lbl 20 `"Yes"', add
label define hiothgove_lbl 21 `"Yes, information"', add
label define hiothgove_lbl 22 `"Yes, but no information"', add
label define hiothgove_lbl 97 `"Unknown-refused"', add
label define hiothgove_lbl 98 `"Unknown-not ascertained"', add
label define hiothgove_lbl 99 `"Unknown-don't know"', add
label values hiothgove hiothgove_lbl

label define histatee_lbl 10 `"No"'
label define histatee_lbl 20 `"Yes"', add
label define histatee_lbl 21 `"Yes, information"', add
label define histatee_lbl 22 `"Yes, but no information"', add
label define histatee_lbl 97 `"Unknown-refused"', add
label define histatee_lbl 98 `"Unknown-not ascertained"', add
label define histatee_lbl 99 `"Unknown-don't know"', add
label values histatee histatee_lbl

label define himcaide_lbl 1 `"No"'
label define himcaide_lbl 2 `"Yes, information"', add
label define himcaide_lbl 3 `"Yes, but no information"', add
label define himcaide_lbl 7 `"Unknown-refused"', add
label define himcaide_lbl 8 `"Unknown-not ascertained"', add
label define himcaide_lbl 9 `"Unknown-don't know"', add
label values himcaide himcaide_lbl

label define himcaree_lbl 0 `"NIU"'
label define himcaree_lbl 1 `"No"', add
label define himcaree_lbl 2 `"Yes, information"', add
label define himcaree_lbl 3 `"Yes, but no information"', add
label define himcaree_lbl 7 `"Unknown-refused"', add
label define himcaree_lbl 8 `"Unknown-not ascertained"', add
label define himcaree_lbl 9 `"Unknown-don't know"', add
label values himcaree himcaree_lbl

label define sdentale_lbl 0 `"NIU"'
label define sdentale_lbl 1 `"Not mentioned"', add
label define sdentale_lbl 2 `"Mentioned"', add
label define sdentale_lbl 7 `"Unknown-refused"', add
label define sdentale_lbl 8 `"Unknown-not ascertained"', add
label define sdentale_lbl 9 `"Unknown-don't ask"', add
label values sdentale sdentale_lbl

label define hinotcov_lbl 00 `"NIU"'
label define hinotcov_lbl 01 `"No, has coverage"', add
label define hinotcov_lbl 02 `"Yes, has no coverage"', add
label define hinotcov_lbl 09 `"Unknown/refused"', add
label values hinotcov hinotcov_lbl

label define hiprivate_lbl 00 `"NIU"'
label define hiprivate_lbl 01 `"No"', add
label define hiprivate_lbl 02 `"Yes"', add
label define hiprivate_lbl 07 `"Unknown-refused"', add
label define hiprivate_lbl 08 `"Unknown-not ascertained"', add
label define hiprivate_lbl 09 `"Unknown-don't know"', add
label values hiprivate hiprivate_lbl

label define hilast_lbl 00 `"NIU"'
label define hilast_lbl 10 `"1 year or less"', add
label define hilast_lbl 11 `"6 months or less"', add
label define hilast_lbl 12 `"More than 6 months, less than 1 year"', add
label define hilast_lbl 13 `"More than 6 months, 1 year or less"', add
label define hilast_lbl 14 `"Less than 1 year"', add
label define hilast_lbl 15 `"1 year"', add
label define hilast_lbl 20 `"1 to 3 years"', add
label define hilast_lbl 21 `"More than 1 year, 3 years or less"', add
label define hilast_lbl 22 `"More than 1 year, less than 3 years"', add
label define hilast_lbl 23 `"1 year or more, less than 2 years"', add
label define hilast_lbl 24 `"2 years or more, less than 3 years"', add
label define hilast_lbl 30 `"3+ years"', add
label define hilast_lbl 31 `"3 years or more"', add
label define hilast_lbl 32 `"More than 3 years"', add
label define hilast_lbl 33 `"3 years or more, less than 5 years"', add
label define hilast_lbl 34 `"5 years or more, less than 10 years"', add
label define hilast_lbl 35 `"10 years or more"', add
label define hilast_lbl 40 `"Never"', add
label define hilast_lbl 97 `"Unknown-refused"', add
label define hilast_lbl 98 `"Unknown-not ascertained"', add
label define hilast_lbl 99 `"Unknown-don't know"', add
label values hilast hilast_lbl

label define hinotyr_lbl 0 `"NIU"'
label define hinotyr_lbl 1 `"No"', add
label define hinotyr_lbl 2 `"Yes"', add
label define hinotyr_lbl 7 `"Unknown-refused"', add
label define hinotyr_lbl 8 `"Unknown-not ascertained"', add
label define hinotyr_lbl 9 `"Unknown-don't know"', add
label values hinotyr hinotyr_lbl

label define hinotyrmo_lbl 00 `"NIU"'
label define hinotyrmo_lbl 01 `"1 month"', add
label define hinotyrmo_lbl 02 `"2 months"', add
label define hinotyrmo_lbl 03 `"3 months"', add
label define hinotyrmo_lbl 04 `"4 months"', add
label define hinotyrmo_lbl 05 `"5 months"', add
label define hinotyrmo_lbl 06 `"6 months"', add
label define hinotyrmo_lbl 07 `"7 months"', add
label define hinotyrmo_lbl 08 `"8 months"', add
label define hinotyrmo_lbl 09 `"9 months"', add
label define hinotyrmo_lbl 10 `"10 months"', add
label define hinotyrmo_lbl 11 `"11 months"', add
label define hinotyrmo_lbl 12 `"12 months"', add
label define hinotyrmo_lbl 97 `"Unknown-refused"', add
label define hinotyrmo_lbl 98 `"Unknown-not ascertained"', add
label define hinotyrmo_lbl 99 `"Unknown-don't know"', add
label values hinotyrmo hinotyrmo_lbl

label define hip1dental_lbl 0 `"NIU"'
label define hip1dental_lbl 1 `"No"', add
label define hip1dental_lbl 2 `"Yes"', add
label define hip1dental_lbl 7 `"Unknown-refused"', add
label define hip1dental_lbl 8 `"Unknown-not ascertained"', add
label define hip1dental_lbl 9 `"Unknown-don't know"', add
label values hip1dental hip1dental_lbl

label define opioidapain_lbl 0 `"NIU"'
label define opioidapain_lbl 1 `"No"', add
label define opioidapain_lbl 2 `"Yes"', add
label define opioidapain_lbl 7 `"Unknown-refused"', add
label define opioidapain_lbl 8 `"Unknown-not ascertained"', add
label define opioidapain_lbl 9 `"Unknown-don't know"', add
label values opioidapain opioidapain_lbl

label define opioidcpain_lbl 0 `"NIU"'
label define opioidcpain_lbl 1 `"No"', add
label define opioidcpain_lbl 2 `"Yes"', add
label define opioidcpain_lbl 7 `"Unknown-refused"', add
label define opioidcpain_lbl 8 `"Unknown-not ascertained"', add
label define opioidcpain_lbl 9 `"Unknown-don't know"', add
label values opioidcpain opioidcpain_lbl

label define dentint_lbl 000 `"NIU"'
label define dentint_lbl 100 `"Never"', add
label define dentint_lbl 200 `"Within the past year"', add
label define dentint_lbl 210 `"Six months or less"', add
label define dentint_lbl 211 `"Visit in Past 2 weeks"', add
label define dentint_lbl 212 `"2 weeks - less than 6 months"', add
label define dentint_lbl 213 `"Greater than 2 weeks - less than 12 months"', add
label define dentint_lbl 220 `"Six months to a year"', add
label define dentint_lbl 221 `"6 months - less than 12 months"', add
label define dentint_lbl 222 `"Greater than 6 months - 12 months"', add
label define dentint_lbl 300 `"1 year - 5 years"', add
label define dentint_lbl 301 `"1 year - less than 2 years"', add
label define dentint_lbl 302 `"Greater than 1 year - 2 years"', add
label define dentint_lbl 303 `"Greater than 1 year - 3 years"', add
label define dentint_lbl 304 `"2 years - less than 3 years"', add
label define dentint_lbl 305 `"2 - 4 years"', add
label define dentint_lbl 306 `"Greater than 2 years - 5 years"', add
label define dentint_lbl 400 `"Greater than 3 years"', add
label define dentint_lbl 401 `"3 years - less than 5 years"', add
label define dentint_lbl 500 `"5+ years"', add
label define dentint_lbl 501 `"Greater than 5 years"', add
label define dentint_lbl 502 `"5 - 9 years"', add
label define dentint_lbl 503 `"10+ years"', add
label define dentint_lbl 990 `"Unknown-all causes"', add
label define dentint_lbl 997 `"Unknown-refused"', add
label define dentint_lbl 998 `"Unknown-not ascertained"', add
label define dentint_lbl 999 `"Unknown-don't know"', add
label values dentint dentint_lbl

label define asthmaev_lbl 0 `"NIU"'
label define asthmaev_lbl 1 `"No or not mentioned"', add
label define asthmaev_lbl 2 `"Yes"', add
label define asthmaev_lbl 7 `"Unknown-refused"', add
label define asthmaev_lbl 8 `"Unknown-not ascertained"', add
label define asthmaev_lbl 9 `"Unknown-don't know"', add
label values asthmaev asthmaev_lbl

label define asthmastil_lbl 0 `"NIU"'
label define asthmastil_lbl 1 `"No"', add
label define asthmastil_lbl 2 `"Yes"', add
label define asthmastil_lbl 7 `"Unknown-refused"', add
label define asthmastil_lbl 8 `"Unknown-not ascertained"', add
label define asthmastil_lbl 9 `"Unknown-don't know"', add
label values asthmastil asthmastil_lbl

label define cancerev_lbl 0 `"NIU"'
label define cancerev_lbl 1 `"No"', add
label define cancerev_lbl 2 `"Yes"', add
label define cancerev_lbl 7 `"Unknown-refused"', add
label define cancerev_lbl 8 `"Unknown-not ascertained"', add
label define cancerev_lbl 9 `"Unknown-don't know"', add
label values cancerev cancerev_lbl

label define cheartdiev_lbl 0 `"NIU"'
label define cheartdiev_lbl 1 `"No"', add
label define cheartdiev_lbl 2 `"Yes"', add
label define cheartdiev_lbl 7 `"Unknown--refused"', add
label define cheartdiev_lbl 8 `"Unknown--not ascertained"', add
label define cheartdiev_lbl 9 `"Unknown--don't know"', add
label values cheartdiev cheartdiev_lbl

label define cholhighev_lbl 0 `"NIU"'
label define cholhighev_lbl 1 `"No"', add
label define cholhighev_lbl 2 `"Yes"', add
label define cholhighev_lbl 7 `"Unknown-refused"', add
label define cholhighev_lbl 8 `"Unknown-not ascertained"', add
label define cholhighev_lbl 9 `"Unknown-don't know"', add
label values cholhighev cholhighev_lbl

label define cholhighyr_lbl 0 `"NIU"'
label define cholhighyr_lbl 1 `"No"', add
label define cholhighyr_lbl 2 `"Yes"', add
label define cholhighyr_lbl 7 `"Unknown-refused"', add
label define cholhighyr_lbl 8 `"Unknown-not ascertained"', add
label define cholhighyr_lbl 9 `"Unknown-don't know"', add
label values cholhighyr cholhighyr_lbl

label define dementiaev_lbl 0 `"NIU"'
label define dementiaev_lbl 1 `"No"', add
label define dementiaev_lbl 2 `"Yes"', add
label define dementiaev_lbl 7 `"Unknown-refused"', add
label define dementiaev_lbl 8 `"Unknown-not ascertained"', add
label define dementiaev_lbl 9 `"Unknown-don't know"', add
label values dementiaev dementiaev_lbl

label define diabeticev_lbl 0 `"NIU"'
label define diabeticev_lbl 1 `"No or not mentioned"', add
label define diabeticev_lbl 2 `"Yes or mentioned"', add
label define diabeticev_lbl 3 `"Borderline"', add
label define diabeticev_lbl 7 `"Unknown-refused"', add
label define diabeticev_lbl 8 `"Unknown-not ascertained"', add
label define diabeticev_lbl 9 `"Unknown-don't know"', add
label values diabeticev diabeticev_lbl

label define heartattev_lbl 0 `"NIU"'
label define heartattev_lbl 1 `"No"', add
label define heartattev_lbl 2 `"Yes"', add
label define heartattev_lbl 7 `"Unknown--refused"', add
label define heartattev_lbl 8 `"Unknown--not ascertained"', add
label define heartattev_lbl 9 `"Unknown--don't know"', add
label values heartattev heartattev_lbl

label define hypertenev_lbl 0 `"NIU"'
label define hypertenev_lbl 1 `"No"', add
label define hypertenev_lbl 2 `"Yes"', add
label define hypertenev_lbl 7 `"Unknown-refused"', add
label define hypertenev_lbl 8 `"Unknown-not ascertained"', add
label define hypertenev_lbl 9 `"Unknown-don't know"', add
label values hypertenev hypertenev_lbl

label define hypertenyr_lbl 0 `"NIU"'
label define hypertenyr_lbl 1 `"No"', add
label define hypertenyr_lbl 2 `"Yes"', add
label define hypertenyr_lbl 7 `"Unknown-refused"', add
label define hypertenyr_lbl 8 `"Unknown-not ascertained"', add
label define hypertenyr_lbl 9 `"Unknown-don't know"', add
label values hypertenyr hypertenyr_lbl

label define strokev_lbl 0 `"NIU"'
label define strokev_lbl 1 `"No"', add
label define strokev_lbl 2 `"Yes"', add
label define strokev_lbl 7 `"Unknown-refused"', add
label define strokev_lbl 8 `"Unknown-not ascertained"', add
label define strokev_lbl 9 `"Unknown-don't know"', add
label values strokev strokev_lbl

label define copdev_lbl 0 `"NIU"'
label define copdev_lbl 1 `"No"', add
label define copdev_lbl 2 `"Yes"', add
label define copdev_lbl 7 `"Unknown-refused"', add
label define copdev_lbl 8 `"Unknown-not ascertained"', add
label define copdev_lbl 9 `"Unknown-don't know"', add
label values copdev copdev_lbl

label define painfreq3mo_lbl 0 `"NIU"'
label define painfreq3mo_lbl 1 `"Never"', add
label define painfreq3mo_lbl 2 `"Some days"', add
label define painfreq3mo_lbl 3 `"Most days"', add
label define painfreq3mo_lbl 4 `"Every day"', add
label define painfreq3mo_lbl 7 `"Unknown-refused"', add
label define painfreq3mo_lbl 8 `"Unknown-not ascertained"', add
label define painfreq3mo_lbl 9 `"Unknown-don't know"', add
label values painfreq3mo painfreq3mo_lbl

label define painfeelevl_lbl 0 `"NIU"'
label define painfeelevl_lbl 1 `"A little"', add
label define painfeelevl_lbl 2 `"A lot"', add
label define painfeelevl_lbl 3 `"Somewhere in between a little and a lot"', add
label define painfeelevl_lbl 7 `"Unknown-refused"', add
label define painfeelevl_lbl 8 `"Unknown-not ascertained"', add
label define painfeelevl_lbl 9 `"Unknown-don't know"', add
label values painfeelevl painfeelevl_lbl

label define paintooth3m_lbl 0 `"NIU"'
label define paintooth3m_lbl 1 `"Not at all"', add
label define paintooth3m_lbl 2 `"A little"', add
label define paintooth3m_lbl 3 `"A lot"', add
label define paintooth3m_lbl 4 `"Somewhere in between a little and a lot"', add
label define paintooth3m_lbl 7 `"Unknown-refused"', add
label define paintooth3m_lbl 8 `"Unknown-not ascertained"', add
label define paintooth3m_lbl 9 `"Unknown-don't know"', add
label values paintooth3m paintooth3m_lbl

label define cvddiag_lbl 0 `"NIU"'
label define cvddiag_lbl 1 `"No"', add
label define cvddiag_lbl 2 `"Yes"', add
label define cvddiag_lbl 7 `"Unknown-refused"', add
label define cvddiag_lbl 8 `"Unknown-not ascertained"', add
label define cvddiag_lbl 9 `"Unknown-don't know"', add
label values cvddiag cvddiag_lbl

label define cvdtest_lbl 0 `"NIU"'
label define cvdtest_lbl 1 `"No"', add
label define cvdtest_lbl 2 `"Yes"', add
label define cvdtest_lbl 7 `"Unknown-refused"', add
label define cvdtest_lbl 8 `"Unknown-not ascertained"', add
label define cvdtest_lbl 9 `"Unknown-don't know"', add
label values cvdtest cvdtest_lbl

label define cvdtestrslt_lbl 0 `"NIU"'
label define cvdtestrslt_lbl 1 `"No"', add
label define cvdtestrslt_lbl 2 `"Yes"', add
label define cvdtestrslt_lbl 3 `"Did not receive results"', add
label define cvdtestrslt_lbl 7 `"Unknown-refused"', add
label define cvdtestrslt_lbl 8 `"Unknown-not ascertained"', add
label define cvdtestrslt_lbl 9 `"Unknown-don't know"', add
label values cvdtestrslt cvdtestrslt_lbl

label define cvdsymp_lbl 0 `"NIU"'
label define cvdsymp_lbl 1 `"No symptoms"', add
label define cvdsymp_lbl 2 `"Mild symptoms"', add
label define cvdsymp_lbl 3 `"Moderate symptoms"', add
label define cvdsymp_lbl 4 `"Severe symptoms"', add
label define cvdsymp_lbl 7 `"Unknown-refused"', add
label define cvdsymp_lbl 8 `"Unknown-not ascertained"', add
label define cvdsymp_lbl 9 `"Unknown-don't know"', add
label values cvdsymp cvdsymp_lbl

label define cvdcanothtr_lbl 0 `"NIU"'
label define cvdcanothtr_lbl 1 `"No"', add
label define cvdcanothtr_lbl 2 `"Yes"', add
label define cvdcanothtr_lbl 7 `"Unknown-refused"', add
label define cvdcanothtr_lbl 8 `"Unknown-not ascertained"', add
label define cvdcanothtr_lbl 9 `"Unknown-don't know"', add
label values cvdcanothtr cvdcanothtr_lbl

label define cvddlycare_lbl 0 `"NIU"'
label define cvddlycare_lbl 1 `"No"', add
label define cvddlycare_lbl 2 `"Yes"', add
label define cvddlycare_lbl 7 `"Unknown-refused"', add
label define cvddlycare_lbl 8 `"Unknown-not ascertained"', add
label define cvddlycare_lbl 9 `"Unknown-don't know"', add
label values cvddlycare cvddlycare_lbl

label define cvddngcare_lbl 0 `"NIU"'
label define cvddngcare_lbl 1 `"No"', add
label define cvddngcare_lbl 2 `"Yes"', add
label define cvddngcare_lbl 7 `"Unknown-refused"', add
label define cvddngcare_lbl 8 `"Unknown-not ascertained"', add
label define cvddngcare_lbl 9 `"Unknown-don't know"', add
label values cvddngcare cvddngcare_lbl

save "C:\Users\jsemprini\OneDrive - University of Iowa\4-Misc Projects\d-NHIS-covid\nhis_raw\2019-2020_nhis_label_050222.dta"
```
