## Supplementary material for "Estimating the within-person change in dental service access measures during the COVID-19 Pandemic": Replication Files and Data Dictionary: nhis_clean_analyze.do

```
clear all

***USE DATA CREATED FROM IPUMS EXTRACT & DO.FILE****

*id 2019-2020 cohort*

gen cohortid=nhispidprvyr if year==2020
replace cohortid=nhispid if year==2019

 duplicates tag cohortid, generate(samplecohort)
 
tab year samplecohort, missing

drop if samplecohort!=1

tab year samplecohort, missing

egen id = group(cohortid)

sum id

order id


****clean potential controls****

gen x_age= age if age<86

gen x_male=0 if sex==2
replace x_male=1 if sex==1

tab marstat if marstat!=99, gen(x_mar) nolab

tab racea 
tab racea , nolab

tab racea if racea!=980, gen(race)

sum race*

gen x_white=.
replace x_white=0 if race1==0 & race6==0
replace x_white=1 if race1==1

tab hispeth, nolab
tab hispeth, gen(hisp)

gen x_nothisp=.
replace x_nothisp=0 if hisp1==2 | hisp2==1
replace x_nothisp=1 if hisp1==1


tab usborn citizen, missing

tab citizen, nolab
tab citizen, gen(cit)

gen x_citizen=0 if cit1==1
replace x_citizen=1 if cit2==1


tab educ, nolab
tab educ, gen(ed)

gen x_hsorless=0 if ed1!=1 & ed2!=1 & ed3!=1 & ed4!=1 & educ<900
replace x_hsorless=1 if ed1==1 | ed2==1 | ed3==1 | ed4==1 

gen x_somecol=0 if ed6!=1 & ed7!=1 & ed8!=1 & educ<900
replace x_somecol=1 if ed6==1 | ed7==1 | ed8==1

gen x_atleastbach=.
replace x_atleastbach=0 if x_hsorless==1 | x_somecol==1
replace x_atleastbach=1 if x_hsorless==0 & x_somecol==0 & educ<900

sum ed*
foreach n of numlist 1/14{
	drop ed`n'
}

tab spouseduc, missing nolab
tab spouseduc, gen(ed)

 gen x_sphsorless=0 if ed1!=1 & ed2!=1 & ed3!=1 & ed4!=1 & educ<900
replace x_sphsorless=1 if ed1==1 | ed2==1 | ed3==1 | ed4==1 

gen x_spsomecol=0 if ed6!=1 & ed7!=1 & ed8!=1 & educ<900
replace x_spsomecol=1 if ed6==1 | ed7==1 | ed8==1

gen x_spatleastbach=.
replace x_spatleastbach=0 if x_sphsorless==1 | x_spsomecol==1
replace x_spatleastbach=1 if x_sphsorless==0 & x_spsomecol==0 & educ<900

tab empstat, nolab
tab empstat

gen x_employed=0 if empstat==200
replace x_employed=1 if empstat==100

tab emplast, nolab

gen x_uethisyr=0 if empstat!=999 & emplast<7
replace x_uethisyr=1 if emplast==1

tab paidsick , nolab

gen x_sickleave=0 if paidsick<7
replace x_sickleave=1 if paidsick==2

tab emphi, nolab
tab emphi 

gen x_work_hi=0 if emphi<7
replace x_work_hi=1 if emphi==2


tab empft, nolab
tab empft

gen x_fulltime=0 if empft<7
replace x_fulltime=1 if empft==2

tab incfam07on, gen(x_hhinc)

tab poverty, gen(x_pov)

*****clean / code dependent and instrument variables (health, insurance)***

foreach x in painfreq3mo painfeelevl paintooth3m{ 
	
	tab year `x', missing
}

tab dentint, nolab
tab dentint

gen y_delaydt=0 if delaydt==1
replace y_delaydt=1 if delaydt==2

gen y_unabledt=0 if unmtnddt==1
replace y_unabledt=1 if unmtnddt==2

tab dentint if dentint<900, gen(lastdent)

sum lastdent*

rename (lastdent*) (y_neverdent y_01yrdent y_12yrdent y_23yrdent y_35yrdent y_59yrdent y_10dent)

tab hip1dental, nolab
tab hip1dental

gen z_dentcov=0 if hip1dental==0 | hip1dental==1
replace z_dentcov=1 if hip1dental==2


****cleat covid/chronic disease variables****
gen g_asthmaev=0 if asthmaev==1
replace g_asthmaev=1 if asthmaev==2

gen g_astmanow=0 if g_asthmaev==0 | asthmastil==1
replace g_astmanow=1 if asthmastil==2

gen g_cancer=0 if cancerev==1
replace g_cancer=1 if cancerev==2

gen g_heartdis=0 if cheartdiev==1
replace g_heartdis=1 if cheartdiev==2

gen g_highcholev=0 if cholhighev==1
replace g_highcholev=1 if cholhighev==2

gen g_highcholyr=0 if g_highcholev==0 | cholhighyr==1
replace g_highcholyr=1 if cholhighyr==2

gen g_diab=0 if diabeticev==1
replace g_diab=1 if diabeticev==2

gen g_heartattackev=0 if heartattev==1
replace g_heartattackev=1 if heartattev==2

gen g_hypertenev=0 if hypertenev==1
replace g_hypertenev=1 if hypertenev==2

gen g_hypertenyr=0 if g_hypertenev==0 | hypertenyr==1
replace g_hypertenyr=1 if hypertenyr==2

gen g_stroke= 0 if strokev==1 
replace g_stroke=1 if strokev==2

gen g_copd=0 if copdev==1
replace g_copd=1 if copdev==2

gen anycdev=0
foreach y in g_asthmaev  g_cancer g_heartdis g_highcholev  g_diab g_heartattackev g_hypertenev  g_stroke g_copd{ 
	
	replace anycdev=1 if `y'==1

}
gen nowcd=0
foreach y in g_astmanow g_highcholyr g_hypertenyr{
	replace nowcd=1 if `y'==1
}

gen totalcdev=g_asthmaev+g_cancer+g_heartdis+g_highcholev+g_diab+g_heartattackev+g_hypertenev+g_stroke+g_copd

rename totalcdev g_totalcddev

gen g_cooccuevr=0 if g_totalcddev<=1
replace g_cooccuevr=1 if g_totalcddev>1

gen g_totalcdnow=g_astmanow+g_highcholyr+g_hypertenyr

gen g_cooccuryr=0 if g_totalcdnow<=1
replace g_cooccuryr=1 if g_totalcdnow>1

keep id year serial quarter strata psu nhishid intervwqtr region urbrrl pernum nhispid nhispidprvyr hhx sampweight longweight partweight x_* h_* y_* t_* z_* g_* c_*  

****SAVE DATA TO ANALYZE****


global visit_outcomes y_delaydt y_unabledt y_neverdent y_01yrdent y_12yrdent y_23yrdent y_35yrdent y_59yrdent  z_dentcov

global controls x_age x_male x_mar1 x_mar2 x_mar3 x_mar4 x_mar5 x_white x_nothisp x_citizen x_hsorless x_somecol x_atleastbach x_sphsorless x_spsomecol x_spatleastbach x_employed x_uethisyr x_sickleave x_work_hi x_fulltime x_hhinc1 x_hhinc2 x_hhinc3 x_hhinc4 x_hhinc5 x_pov1 x_pov2 x_pov3 x_pov4 x_pov5 x_pov6 x_pov7 x_pov8 x_pov9 x_pov10 x_pov11 x_pov12 x_pov13 x_pov14


sum $outcomes 
sum $controls

tab quarter, nolab
tab region
tab region, nolab
tab urbrrl
tab urbrrl, nolab
tab year

tab region , gen(geo)
tab urbrrl, gen(metro)

***carryforward weight to 2019 observations****
gen cf=0 if year==2020
replace cf=1 if year==2019
xtset id cf

****Requires: ssc install carryforward*****
bysort id: carryforward longweight, gen(finalwt)

foreach c in c_coviddiag c_covidtest c_covidpostest c_symp1 c_symp2 c_symp3 c_symp4 c_symp5 c_delaycare c_missedneededcare{
	
	bysort id: carryforward `c', gen(`c'_full)
}

****create four groups of dental insurance: 00, 01, 10, 11*****
bysort id: carryforward z_dentcov, gen(di_20)

xtset id year

bysort id: carryforward z_dentcov, gen(di_19)

tab di_19 di_20, missing

replace di_20=0 if di_20==.
replace di_19=0 if di_19==.

gen di_cat=.
replace di_cat=1 if di_19==0 & di_20==0
replace di_cat=2 if di_19==1 & di_20==0
replace di_cat=3 if di_19==0 & di_20==1
replace di_cat=4 if di_19==1 & di_20==1

* 1 - never (0,0), 2- early only (1,0), 3 - late only (0,1), 4 - both (1,1)


*** 4% decline in annual visit, 1% increase in pain4****
estimates clear
foreach y in $visit_outcomes {
	
	eststo: xtreg `y' i.year [pw=finalwt], vce(cluster id) fe
}

esttab using test1.csv, b(3) se(3) replace

*** by dental health insurance groups*****
estimates clear
foreach y in  y_delaydt y_unabledt y_neverdent y_01yrdent y_12yrdent {
	foreach n of numlist 1/4{
	
	eststo: xtreg `y' i.year [pw=finalwt] if di_cat==`n', vce(cluster id) fe
}
}

esttab using test2.csv, b(3) se(3) replace


*****design subgroups: 1) risk ; 2) region/rural****

gen cat1=1 if region==1
gen cat2=1 if region==2 
gen cat3=1 if region==3
gen cat4=1 if region==4
gen cat5=1 if urbrrl==1
gen cat6=1 if urbrrl==2
gen cat7=1 if urbrrl==3
gen cat8=1 if urbrrl==4
gen cat9=1 if g_totalcddev==0
gen cat10=1 if g_totalcddev>0
gen cat11=1 if g_cooccuevr==0
gen cat12=1 if g_cooccuevr>0
gen cat13=1 if x_age<40
gen cat14=1 if x_age>=40 & x_age<65
gen cat15=1 if x_age>=65 & x_age<75
gen cat16=1 if x_age>=75


estimates clear
foreach n of numlist 1/16{
foreach y in $visit_outcomes {
	
	eststo: xtreg `y' i.year if cat`n'==1 [pw=finalwt], vce(cluster id) fe
}


}
esttab using test3.csv, b(3) se(3) replace
estimates clear
```
